# MALDI-ToF Protein Profiling as Potential Rapid Diagnostic Platform for COVID-19

**DOI:** 10.1101/2021.05.26.21257798

**Authors:** Prajkta Chivte, Zane LaCasse, Venkata Devesh R. Seethi, Pratool Bharti, Joshua Bland, Shrihari S. Kadkol, Elizabeth R. Gaillard

**Author notes:** co-first authors (Prajkta Chivte, Zane LaCasse).

## Abstract

More than a year after the COVID-19 pandemic has been declared, the need still exists for accurate, rapid, inexpensive and non-invasive diagnostic methods that yield high specificity and sensitivity towards the current and newly emerging SARS-CoV-2 strains. Several studies have since established saliva as a more amenable specimen type for early detection of SARS-CoV-2 as compared to nasopharyngeal swabs. Considering the limitations and high demand for COVID-19 testing, we employed MALDI-ToF mass spectrometry for the analysis of 60 gargle samples from human donors and compared the spectra with their COVID-19 status. Several standards including isolated human serum immunoglobulins and controls such as pre-COVID-19 saliva and heat inactivated SARS-CoV-2 virus were simultaneously analyzed to provide a relative view of the saliva and viral proteome as they would appear in this works methodology. Five potential biomarker peaks were established that demonstrated high concordance with COVID-19 positive individuals. Overall, the agreement of these results with RT-qPCR testing on NP swabs was no less than 90% for the studied cohort, which consisted of young and largely asymptomatic student athletes. From a clinical standpoint, the results from this pilot study are promising and suggest that MALDI-ToF can be used to develop a relatively rapid and inexpensive COVID-19 assay.

## 1. Introduction

Coronavirus disease 2019 (COVID-19), a highly transmissible disease caused by the Severe Acute Respiratory Syndrome Coronavirus 2 (SARS-CoV-2) virus, was discovered in the Hubei Province of China in December 2019 and was soon declared a pandemic by the World Health Organization (WHO) in March 2020. Globally, there have been more than 189 million confirmed cases of COVID-19, including over 4 million deaths, as of July 2021 [1]. Clinical manifestations of COVID-19 predominantly include fever, dry cough, muscle pain, anosmia and fatigue [2]. Infections may also result in a number of medical complications, which worsen in patients of advanced age or with co-morbidities. In contrast to many other viral infections, 48% of persons who test positive for COVID-19 are asymptomatic [3]. Additionally, a report by Johannson et al. [4] concludes that at least 50% of new infections originate from contact with asymptomatic carriers. These reports are difficult to interpret, however, because not all persons who are asymptomatic continue to be asymptomatic throughout the course of infection; percentages of persons who remain asymptomatic are reported to range from 20% - 50% [5],[6].

Facing the ongoing pandemic caused by SARS-CoV-2, early diagnosis of COVID-19 is of great importance to control outbreaks in communities. Thus, it is crucial to design rapid, sensitive and accurate diagnostic tests to address this massive public health crisis. Broadly, there are two major categories of diagnostic tests to detect SARS-CoV-2: i) Detection of the viral RNA genome or ii) detection of viral proteins and antibody response against the virus [7].

The first type of test reports whether the virus is present at an abundance above the analytical sensitivity of the assay. The second type of test detects the presence of antibodies (mostly IgM and IgG) against SARS-CoV-2 within an individual who has been exposed to the virus. The strong antibody responses against the spike (S) and the nucleocapsid (N) proteins have been demonstrated to be of high diagnostic utility [8].

SARS-CoV-2 is an enveloped virus that encodes at least 29 proteins in its RNA genome, four of which are structural proteins: spike (S), membrane (M), envelope (E) and nucleocapsid (N) proteins. The S protein is responsible for binding to the cellular surface receptor angiotensin-converting enzyme 2 (ACE2) through the receptor binding domain (RBD), an essential step for membrane fusion. Activation of the S protein requires cleavage between the S1 and S2 subunits by a furin-like protease and a subsequent conformational change [9].

The current gold standard test for COVID-19 is of the first type of diagnostic test mentioned: molecular testing for the presence of SARS-CoV-2 RNA, which is accomplished by reverse transcription polymerase chain reaction (RT-qPCR). The sensitivity and specificity of RT-qPCR is high depending on the primer/probe sets used. False positive results, although uncommon, can occur because of contamination due to the template amplification nature of RT-qPCR, high viral loads in samples and widespread prevalence of infection. False negative results are generally related to sampling issues, nucleic acid degradation, testing in the very early phase soon after exposure or late convalescent phase of the infection or infections with variants that contain mutations in the primer/probe binding sites. RT-qPCR tests are usually carried out using specimens collected via a nasopharyngeal or oropharyngeal swab or saliva [7], [10], [11].

SARS-CoV-2 and the biomarkers of COVID-19 can be found in multiple other specimen types, however. These include tracheal aspirate, sputum, whole blood, plasma and serum [8]. Of particular interest in finding alternatives to nasopharyngeal swabs, it has been reported that standardized saliva collection can be adopted to detect SARS-CoV-2 infection, as saliva droplets are a main vehicle of viral transmission and the salivary glands are reported to be a target of infection as well as a reservoir for the virus [12],[13]. In fact, approval for using saliva as a primary test material for SARS-CoV-2 was first given to Rutgers’ RUCDR Infinite Biologics and collaborators under emergency use authorization (EUA) by the Food and Drug Administration (FDA) [12]. Similar saliva-based tests were authorized for Yale School of Public Health and implemented by University of Illinois Urbana Champaign [14],[15].

The reliability of viral detection in saliva has been shown with SARS-CoV, Zika and Ebola [16]. Saliva as a specimen for diagnosis provides benefits like rapid, non-invasive collection, cost-effectiveness and patient acceptance. Perhaps of highest diagnostic importance, however, the SARS-CoV-2 load in saliva is reported to be maximum within the first week of symptom onset, providing an avenue for early diagnosis of COVID-19 [17].

In addition to the type of specimen collected for a diagnostic test, it is also crucial to decide upon the type of biomolecule to be detected. Molecular diagnosis of COVID-19 primarily depends on detection of viral RNA. However, RNA is extremely sensitive to degradation by ribonucleases and its extraction process is time-consuming, expensive and demands trained personnel as improper storage and extraction can be a major factor in contributing to false negative COVID-19 results [11]. Additionally, SARS-CoV-2 has mutated over time [18]. This might prove to be a major challenge for tests that operate by detection of viral RNA by RT-qPCR. Mutations in the primer or probe binding sites may decrease or severely limit the ability of the assay to detect SARS-CoV-2. As such, either multiplex RT-qPCR with two or three primer/probe sets may be needed to avoid false negative results. Moreover, a constant and long-term surveillance for mutations in the primer/probe binding sites is needed to ensure adequate test performance.

On the other hand, proteins are more stable molecules that are present in higher amounts in the virus. Proteins cannot be directly amplified like nucleic acids which considerably reduces the chances of producing false positive results due to contamination. Furthermore, several different proteins can be detected in the same analysis which increases the number of diagnostic markers and improves the conclusions drawn from a test. For instance, various classes of proteins like antibodies, cytokines and viral proteins can be collectively analyzed using mass spectrometry and other proteomic techniques for their characterization and quantitation [11],[19]. Beyond merely detecting a single target of interest such as a specific SARS-CoV-2 protein, proteomics combined with informatics can derive actionable information from large data sets and thus help in establishing biomarkers for this disease [20]. Such holistic information is necessary to understand the pathogenesis of SARS-CoV-2 infection and may lead to novel therapies not only against the virus, but also help modify the host response to the virus.

Developing a diagnostic test that overcomes the limitations of nucleic acid testing would assist in fulfilling the worldwide demand of reliable, rapid, accurate and cost-effective testing. Iles et al. have published a preliminary report on utilizing water gargle samples to monitor COVID-19 associated proteins using MALDI-ToF mass spectrometry [21]. The current study proceeds in a similar direction and utilizes saliva for testing SARS-CoV-2 through a simple gargle procedure followed by MALDI-ToF mass spectrometric analysis. However, in this study, the mass spectral analysis was compared to the RT-qPCR status of the individuals from NP swabs. The method described in this article potentially detects both viral proteins and the antibodies produced against them. For the 60 saliva samples that were analyzed, the area under the curve (AUC) of potential viral protein and host protein peaks was used to detect the presence of SARS-CoV-2 and the host immune response against the virus. The values for AUC of the biomarkers and viral proteins were compared to RT-qPCR results from nasopharyngeal swabs sampled in parallel.

## 2. Materials and Methods

### 2.1 Ethical and biosafety statement

This work was approved by the Institutional Review Board and Institutional Biosafety Committee of Northern Illinois University (NIU) (August 12, 2020) as well as the Institutional Review Board of the University of Illinois at Chicago (February 11, 2021). Informed consent was obtained with the signature of the volunteers. Personal identification was not associated with any sample and collected information was limited to demographics, symptoms, and RT-qPCR results. Sample handling and processing followed all biosafety level guidelines.

### 2.2 Sample collection

Samples were collected from a drive-thru gateway testing program for student athletes organized by NIU Athletics at the Yordon Center in DeKalb, Illinois between the months of August and September of 2020. Student athletes received a nasopharyngeal (NP) swab sample collection that was subjected to RT-qPCR testing through the DeKalb County Health Department. NP swabs were collected from individuals and analyzed for SARS-CoV-2 by RT-qPCR with the Abbott RealTime SARS-CoV-2 Assay (EUA). The analysis was done by the University of Illinois Hospital laboratory in Chicago. Results were reported as Detected, Not detected or Inconclusive. Students who consented to participate in the research study were asked to provide a water gargle sample at the same time as the NP swab sample collection. Subjects were asked to gargle 10 mL of bottled spring water for 30 seconds which was then deposited in a 50 mL conical centrifuge tube. Samples were stored at −20 °C until processing and analysis. A total of 550 gargle samples were collected to ensure a substantial pool of COVID-19 positive samples were included. For the current analysis, 60 (30 COVID-19 positives + 30 COVID-19 negatives) of the above total samples were processed and analyzed.

### 2.3 Preparation of samples and controls

#### Gargle samples

The gargle samples were prepared and analyzed following the method reported by Iles et al. [21] with minor modifications. Gargle samples were thawed and transferred to a 30 mL disposable polypropylene beaker (Fisher Scientific, Waltham, MA). Approximately 5 mL of each sample was filtered through a 0.45 µm polyethersulfone membrane filter (Fischer Scientific, Waltham, MA) with the filtrate being collected in the original 50 mL tube. Next, acetone precipitation was conducted on the filtrate by adding 5 mL of chilled acetone (Sigma-Aldrich, St. Louis, MO) to each tube. Samples were centrifuged in a Beckman Coulter Avanti J-E series centrifuge with a JA-20 rotor at 16,000 x g for 30 minutes at 4°C. The supernatant was disposed, the rim of the tube patted dry, and the pellet was resuspended using 100 µL of 1 M DTT (Sigma-Aldrich, St. Louis, MO) in LBSD-X buffer (MAPSciences, Bedford, UK), a proprietary membrane dissolution and viral envelope protein solubilization buffer. This buffer (reconstitution buffer) was prepared fresh daily by adding appropriate amounts of DTT solution before each analysis. Finally, to recover as much pelleted material as possible, the 100 µL reconstitution buffer was washed down the sides of the tube multiple times thoroughly. Upon complete reconstitution, the samples were gently vortexed and incubated at room temperature for 15 minutes.

#### Saliva sample for limit of detection (LoD)

SARS-CoV-2 viral load was quantitated by RT-qPCR in saliva samples by using S gene primers and probe as reported previously [22]. The remaining aliquot was frozen at −20□ (or kept on dry ice during transportation) until MALDI-ToF analysis. Then, 500 μL of the specimen was added to 1.5 mL LC-MS grade H_2_O (OmniSolv, Sigma-Aldrich, St. Louis, MO) and this was subjected to 3X v/v chilled acetone (6 mL) precipitation and incubated overnight at −20□. The sample was then processed and analyzed following the gargle sample protocol.

#### Standards and controls

Human serum antibodies IgA (I4036), IgG (I4506) and IgM (I8260) (Sigma-Aldrich, St. Louis, MO) were reduced with 1 M DTT for 10 minutes for a final concentration of 3 pmol of each antibody on the plate. Human salivary α-amylase (A1031, Sigma-Aldrich, St. Louis, MO) was prepared at 100 pmol/μl in LC-MS grade H_2_O, 1 μl of which was spotted. Preparation of positive and negative controls for MALDI-ToF analysis followed closely to that of gargle samples. The negative control, consisting of pooled human saliva (pre-COVID-19) collected before November 2019 (Lee Biosolutions, Maryland Heights, MO), was prepared by spiking 500 µL of the thawed stock to 10 mL of water in a 50 mL tube. From here, the control was filtered and processed following the gargle sample procedure. The positive control (heat inactivated SARS-CoV-2) was obtained from BEI Resources (NR-52286). Briefly, this standard was a heat inactivated, clarified and diluted cell lysate and supernatant from Vero E6 cells infected with SARS-CoV-2. This sample (225 µL) was treated with 4X v/v of chilled acetone (900 µL) and incubated overnight at −20°C followed by centrifugation at 10,000 x g for 15 minutes at 4°C. The pellet was completely dissolved in 25 µL of the reconstitution buffer.

#### Spotting

For the assay, a sandwich method of matrix-sample-matrix spotting was employed. Sinapinic acid (Sigma-Aldrich, St. Louis, MO) was used as the matrix and consisted of 20 mg/mL in a 50:50 LC-MS grade H_2_O to acetonitrile (Oakwood Chemical, Estill, SC) solution containing 0.1% trifluoroacetic acid (Sigma-Aldrich, St. Louis, MO). First, 1 µL of sinapinic acid matrix was spotted in three wells of a 384 well stainless steel MALDI-MS sample plate (Shimadzu, Kyoto, Japan). After this air-dried, 1 µL of sample, control, or standard was spotted in each well immediately followed by 1 µL of sinapinic acid matrix. The matrix was made fresh every 7 days and stored at 4°C between analyses.

### 2.4 Data Acquisition

Spectra acquisition was performed with a Shimadzu AXIMA Performance MALDI-ToF mass spectrometer (Shimadzu Kratos Analytical, Manchester, UK) equipped with a nitrogen laser set at 337.1 nm with a pulse width of 3 ns and maximum repetition rate of 60 Hz.

#### Instrument parameters

The AXIMA Performance mass spectrometer was operated with the Shimadzu Biotech Launchpad Software (version 2.9.4) and was run in positive-ion linear detection mode. The laser power and repetition rate were set at 100 µJ/pulse and 50 Hz respectively. Spectra were acquired by summing 5,000 spectra (250 profiles by 20 shots) in a range of 2,000 to 200,000 m/z per sample by shooting in a raster pattern over the target well. The ion gate was set to blank values below 1,500 m/z. Pulsed extraction was set to 50,000 m/z.

#### Instrument calibration

The instrument was calibrated daily using the (M+H)^+^ and (M+2H)^2+^ peaks of ProteoMass Apomyoglobin MALDI-MS Standard (Sigma-Aldrich, St. Louis, MS), prepared at 100 pmol/µL in LC-MS grade H_2_O and spotted as described above. Signal intensities of the calibrant were recorded throughout the entire analysis to track inter-day instrument performance. Calibration was accepted if the mass deviation was less than 500 mDa.

### 2.5 Data Analysis

Shimadzu Biotech Launchpad was used to export a text file for each gargle sample that was the spectrum of mass-to-charge values ranging from 2,000 m/z to 200,000 m/z alongside respective ion count intensities. In preprocessing, seven subranges (shown in Table 1) were identified where m/z values were presumably indicative of host immune proteins or viral proteins. The intermittent values in each of the seven subranges indicated the presence of similar protein masses, hence ion counts in each subrange were coalesced together through integration of points to calculate the area under curve (AUC) which produced seven features for each data sample. AUC was computed by leveraging the composite Simpson’s rule for each spectral range [23]. Simpson’s rule is a numerical integration method that divides the integral range into shorter subintervals (indicated by three consecutive data points) and uses a quadratic polynomial on each subinterval to approximate the curve. If we define the width between each point as (*h*), the start and end of each range are *x*_0_ and *x*_*n*_ respectively, where *n* is the total number of points, the width of each integral (*h*) can be calculated as shown in equation (1). One limitation of this approach is the requirement that *h* be uniform between each consecutive point. In the mass spectrometric data, protein peaks had two floating points precision which were unevenly spaced with different widths between two consecutive points. To make the widths uniform, each protein mass was rounded to the nearest integer and the mean of ion counts was taken at each integer value. This made the widths uniform with *h* = 1. If the points on the curve x_0_, x_1_, x_2_, …, x_n_ have corresponding values *f*(x_0_), f(x_1_), *f*(*x*_2_), …, *f*(*x*_*n*_) respectively, then the area for the first subinterval A_1_ can be calculated using equation (2). Summing the areas for all subintervals i.e.,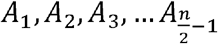 gives the composite function to calculate AUC as shown in the equation (3). Since each curve was not smooth, reduced error can be achieved with Simpson’s rule for integration as compared to trapezoidal rule, which approximates the integral by forming trapezoids with straight lines in each subinterval [24].

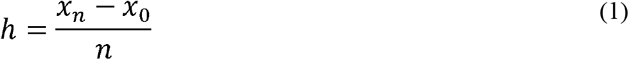

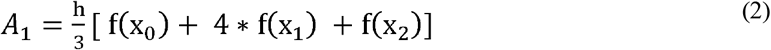

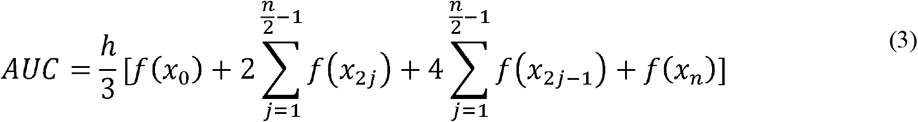

**Table 1:** Potential protein biomarker ranges identified by MALDI-ToF from gargle samples.

| Index | Range (m/z) |
| --- | --- |
| 1 | 11,140 – 11,160 |
| 2 | 23,550 – 23,800 |
| 3 | 27,900 – 29,400 |
| 4 | 55,500 – 59,000 |
| 5 | 66,400 – 68,100 |
| 6 | 78,600 – 80,500 |
| 7 | 111,500 – 115,500 |

## 3. Results and Discussion

Recently, studies utilizing MALDI-MS as a potential method for the detection of SARS-CoV-2 have been reported [25],[26]. However, these studies involved nasopharyngeal swab sampling, were limited to the mass-to-charge range in collected mass spectra (less than 20,000 m/z) and lacked association of peaks to any molecular identity. In another study, published recently by Iles et al. [21] MALDI-MS was employed to detect both viral and human proteins from samples of gargled water. Though this study had a non-invasive sample collection and a wider mass-to-charge range, no direct comparison was made with RT-PCR results in clinical samples.

Herein, we report the analysis of 60 water gargle samples using a MALDI-ToF methodology compared to the RT-qPCR status of individuals done on NP swabs. We did not directly analyze the gargle samples by RT-qPCR and instead relied on the results from NP swabs to classify an individual as positive or negative for SARS-CoV-2. By comparing the mass spectra in known COVID-19 positive and COVID-19 negative individuals, clear distinctions were observed in the range of 20,000 m/z to 200,000 m/z, a range not analyzed by previous COVID-19 MS studies. Furthermore, the area under the curve (AUC) of putative host and viral protein peaks was used to correlate the MALDI-ToF profiles with the RT-qPCR status.

### 3.1. MALDI-ToF profiling in COVID-19 negative individuals

A baseline spectrum of peaks was determined in saliva collected prior to the emergence of COVID-19 (Section 2.3). This spectrum serves as a control against which any changes in COVID-19 positive individuals could be compared. Figure 1 presents the profile for a representative COVID-19 negative sample that is nearly identical in terms of m/z peaks to the spectrum for the negative control. Of particular note are the peaks consistent between the two profiles, for salivary proteins would be present in both diseased and healthy individuals. The identity of these proteins is suggested later (Section 3.3).

**Figure 1.**
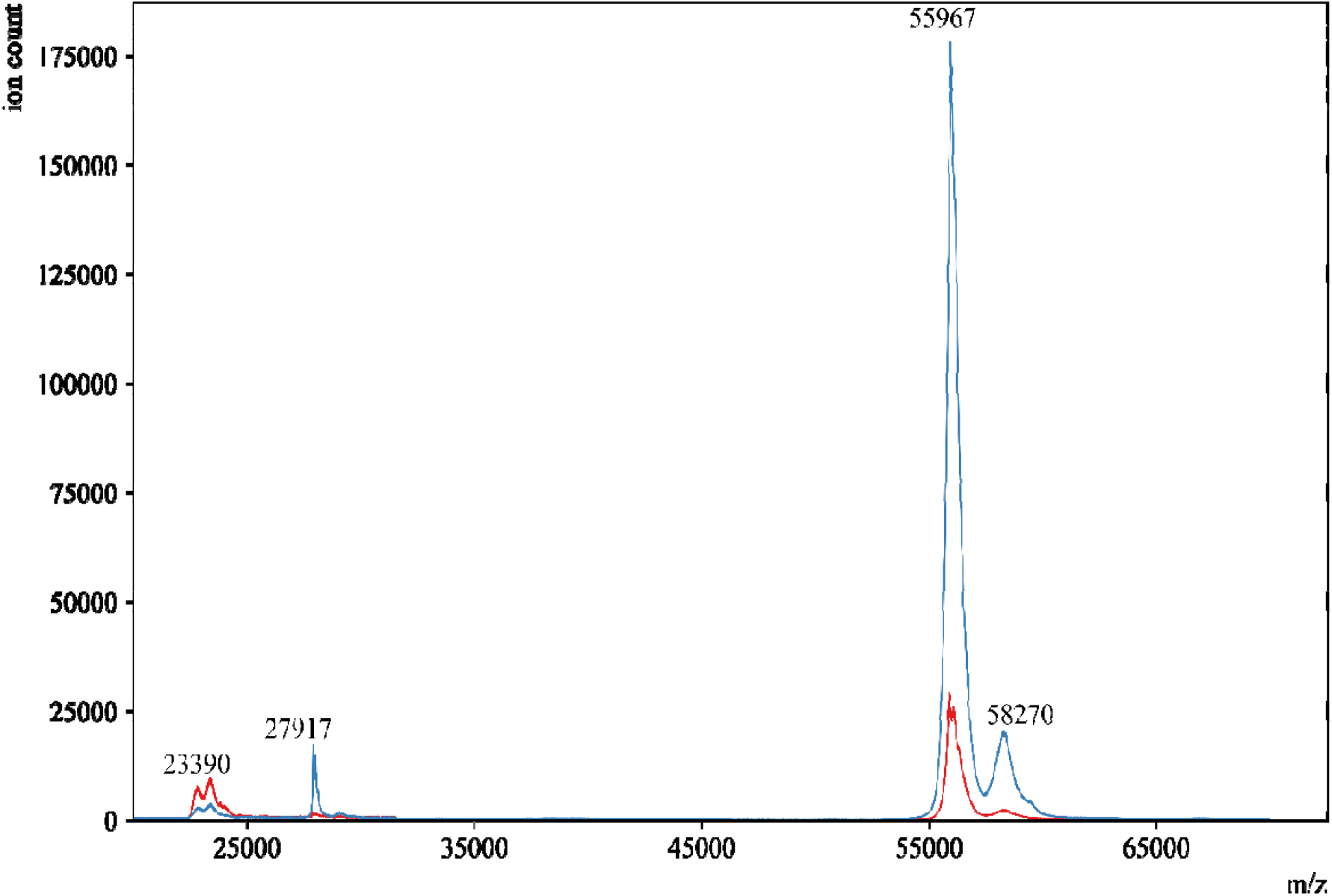
displays the MALDI-ToF mass spectra of a gargle sample from an example donor who tested COVID-19 negative (— red) along with the negative control (—blue). Both profiles closely overlapped each other, and no other signal was detected after 65,000 m/z.

### 3.2. Potential biomarkers for COVID-19 by MALDI-ToF

Distinct differences in MALDI-ToF spectra were observed in individuals who were COVID-19 negative or positive in NP swabs (Figure 2). Overall, gargle samples from COVID-19 positive individuals showed higher intensities along with additional peaks in the spectrum. Peaks around 23,000 m/z, 28,000 m/z and 56,000 m/z were present in both positive and negative individuals. Interestingly, these peaks showed a higher intensity in the COVID-19 positive cases (Figure 2A). Additional peaks that were unique to COVID-19 positive individuals were present between the ranges of 33,000 m/z to 51,000 m/z and 65,000 m/z to 120,000 m/z (Figure 2B and 2C).

**Figure 2.**
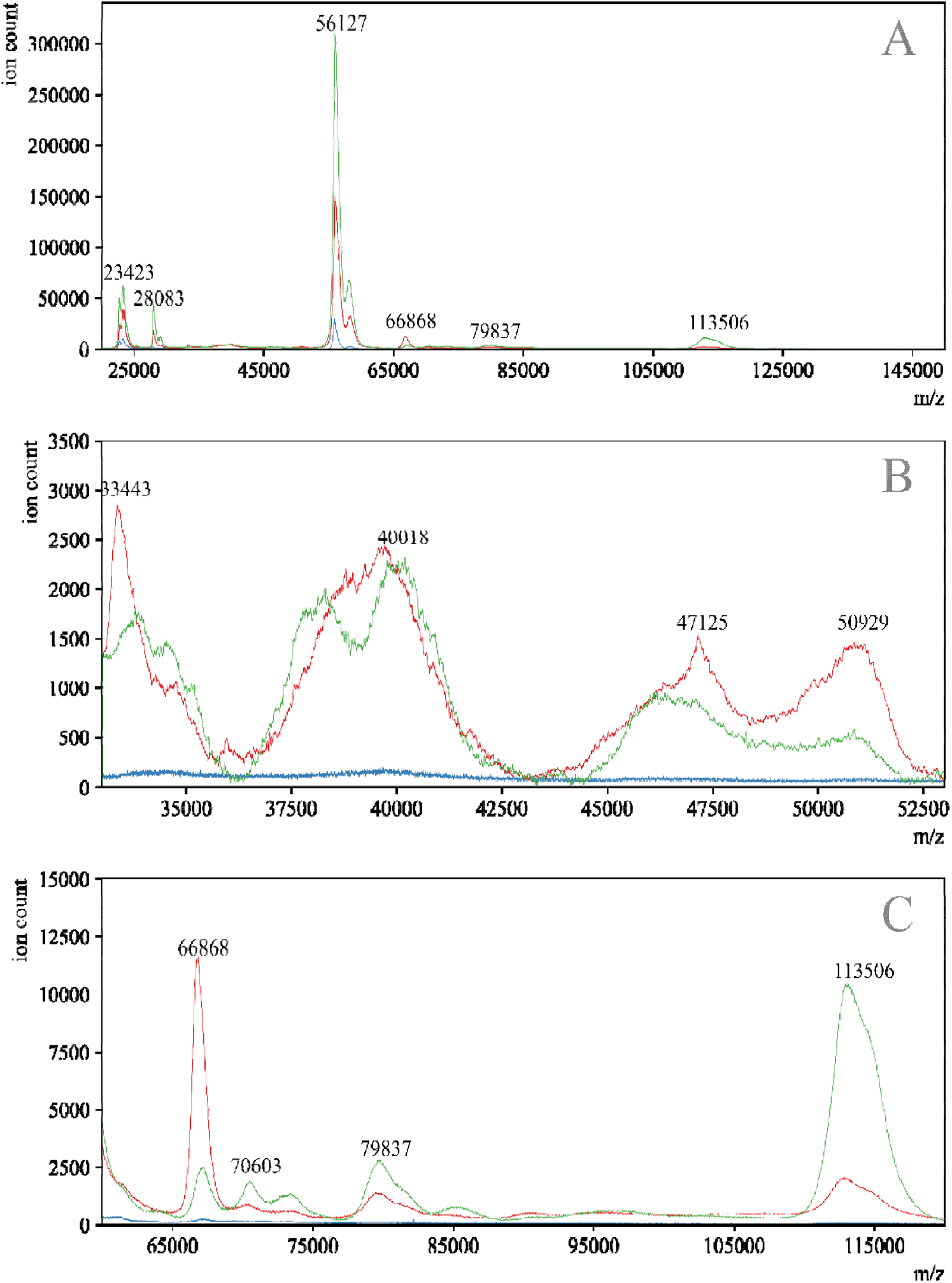
displays the MALDI-ToF mass spectra of a gargle sample from a COVID-19 negative donor (—blue) and an overlay of two COVID-19 positive donors (— red, — green). Panel **A** is the full-range mass spectra of the samples. Panels **B** and **C** are specific ranges where differences in mass spectra are prominently observed between COVID-19 positive and COVID-19 negative samples.

It is important to note that although the peaks found within these ranges reasonably separated the COVID-19 positive from negative individuals, variation in peak intensities was apparent between one COVID-19 positive spectrum and another. We suspect that this may be due to sampling at different time points in the course of the infection and varying immune response in the hosts. Included among the viral and immune proteins discussed is a peak located at 11,150 m/z used as a quality control feature, most likely to be cystatin A, a resident protein of saliva [19]. The presence of this peak was used as an internal control to deem a sample as successfully gargled and appropriate to be included for analysis.

### 3.3. Comparison and identification of peaks in gargle samples

The mass spectra of gargle specimens contain signals from host salivary proteins and viral proteins in COVID-19 positive individuals. Hence, it is crucial to identify peaks that confirm the presence of SARS-CoV-2 in COVID-19 positive individuals along with markers of an immune response against the virus.

To begin, we analyzed the mass spectrum from pre-COVID-19 saliva and a gargle sample from an individual who tested negative in an NP swab by RT-qPCR (Figure 1) to establish a baseline spectrum against which spectra from COVID-19 positive individuals could be compared. The most prominent peak in these spectra was observed near 56,000 m/z. Two smaller peaks are also present near 23,300 m/z and 28,000 m/z. Based on a previous study, we suspected that the peaks near 56,000 m/z and 23,000 m/z most likely represent IgA heavy chain and Ig light chains respectively [21].

To support this notion, we analyzed three human serum immunoglobulins (IgA, IgG and IgM) under reducing conditions using the same protocol as in gargle samples (Figure 3). The heavy chains for IgA, IgG and IgM were detected at 57,372 m/z, 51,142 m/z and 71,678 m/z respectively, while the light chains for these antibodies were found between 23,000 m/z and 24,000 m/z. These results suggest that the peaks around 56,000 m/z and 23,000 m/z in pre-COVID-19 saliva and the gargle samples from a COVID-19 negative individual are indeed IgA heavy chain and Ig light chains. This is not surprising because basal levels of secretory IgA and light chains are nearly always detected in saliva [17]. Various other peaks were observed throughout the spectra of the standards which may represent combinations of heavy and light chains, dimers of heavy chains, and multiply-charged ions.

**Figure 3.**
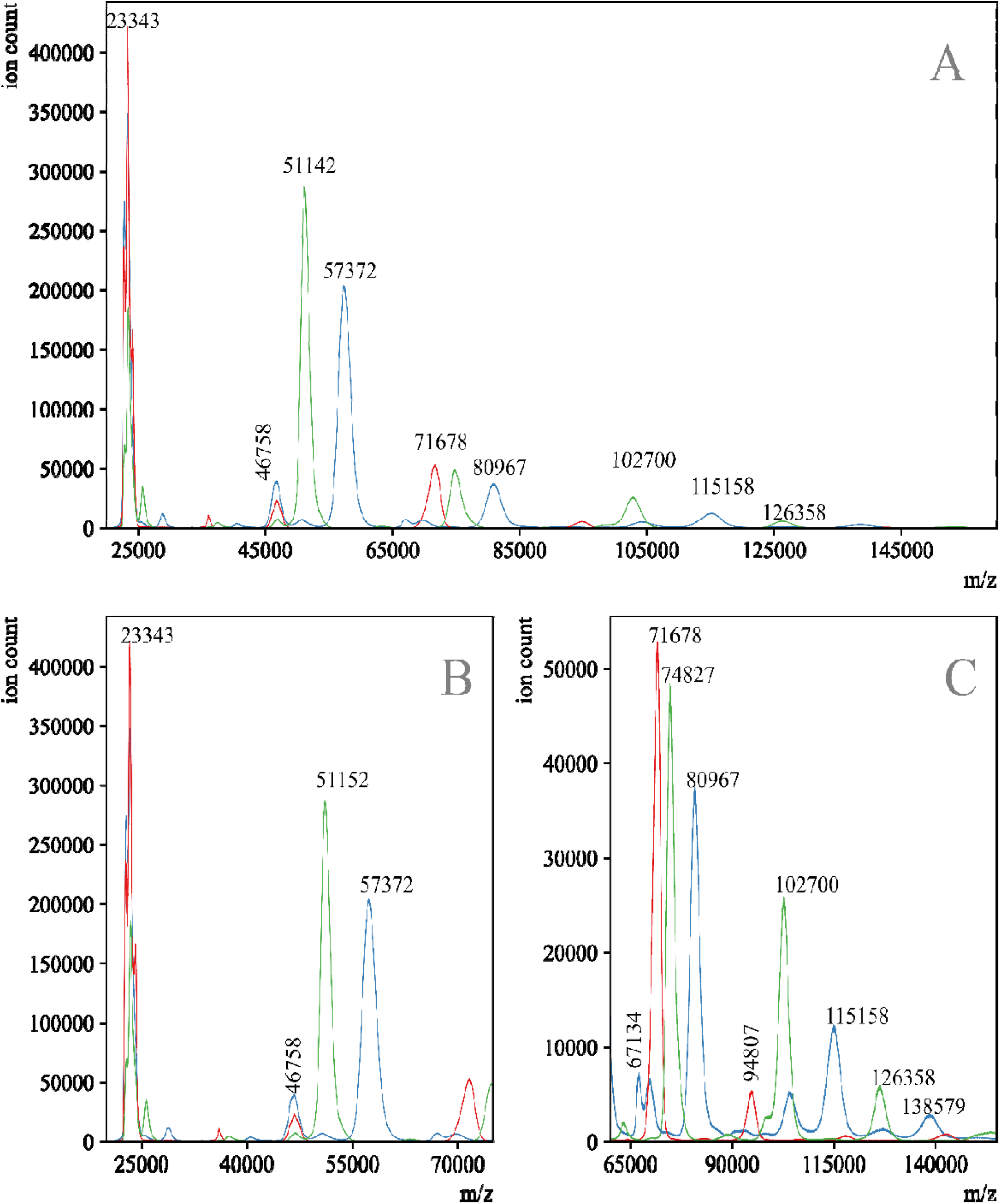
displays the MALDI mass spectra of human serum derived IgA (—blue), IgG(— green) and IgM (— red) isolated from human serum reduced with 1M DTT for 10 minutes. Panel **A** shows the entire mass range collected and Panels **B** and **C** show zoomed-in ranges.

Interestingly, the IgA heavy chain and Ig light chain peaks were almost twice as intense in COVID-19 positive individuals when compared to COVID-19 negative individuals. We suspect that this may be due to a robust immune response against SARS-CoV-2 upon exposure to the virus. Along the same lines, another feature to note is the existence of a peak at 70,603 m/z in COVID-19 positive individuals that could correspond to the heavy chain peak of IgM at 71,678 m/z. This suggests an early response to infection in COVID-19 positive individuals, as it has been reported that viral-specific IgM antibodies are produced first followed by IgA and IgG [27].

Additionally, human α-amylase is found in two forms in saliva with molecular weights of around 56 kDa (unglycosylated) and 62 kDa (glycosylated) [28],[29]. MALDI-MS data have also been reported for human salivary α-amylase and a Y151M mutant, both having a parent ion peak near 56 kDa [30],[31]. Because these masses fall within the region discussed above regarding heavy chains of IgA, samples of α-amylase were also analyzed by MALDI-MS. The observed spectra exhibit a peak at 56,600 m/z with a shoulder at 58,000 m/z (Figure S1), therefore α-amylase overlaps with the IgA heavy chain peak in the gargle sample spectra.

Apart from the IgM heavy chain peak at 71,000 m/z, additional peaks were observed between 65,000 m/z and 120,000 m/z in gargle samples from COVID-19 positive individuals. Peaks were prominent in the ranges of 66,400 to 68,100 m/z and 78,600 to 80,500 m/z (Figure 2C). Comparing the UniProt database (UniProtKB P0DTC2) and the work of Iles et al., the peak at 79,837 m/z most likely represents a signal for the S1 fragment of the SARS-CoV-2 spike protein; a peak with a similar m/z was also observed in the positive control spectrum from SARS-CoV-2 (Figure 4). Furthermore, the peaks described by Iles et al.[21] as viral envelope proteins (VEPs) were also visible in COVID-19 positive profiles (Figure 2B).

**Figure 4.**
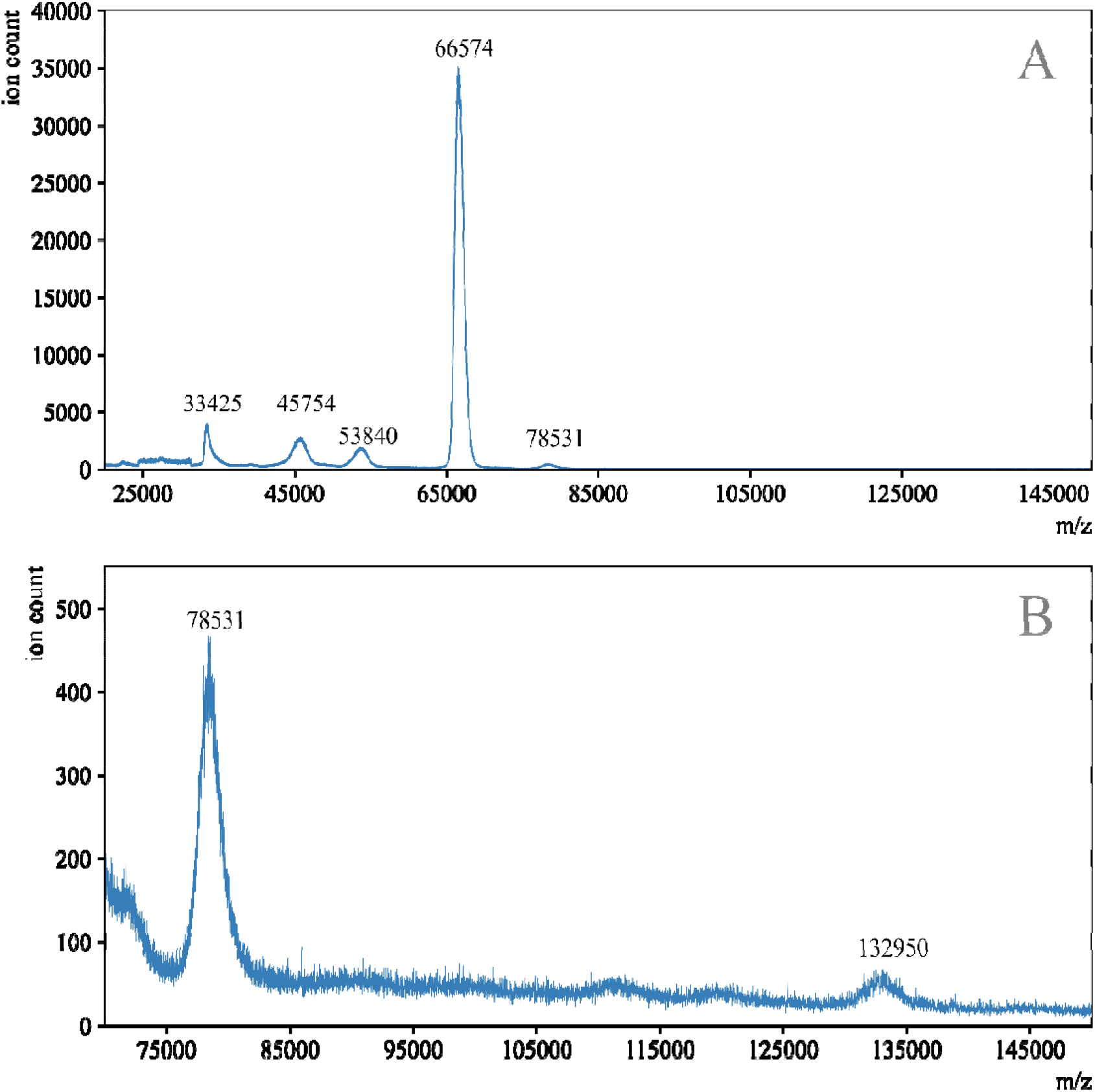
displays the MALDI-ToF spectrum of heat inactivated cell lysate and supernatant of Vero E6 cells infected with SARS-CoV-2, which was utilized as the positive control. Panel **A** shows the entire mass range collected and Panel **B** shows the major peak signals beyond 70,000 m/z.

Compared to COVID-19 negative individuals, a signal was consistently observed in the range of 66,000 m/z to 68,000 m/z in gargle samples from COVID-19 positive individuals. By comparing numerous positive spectra, it appears that there are at least two distinct species with closely overlapping m/z envelopes. This peak could represent the S2 fragment of the SARS-CoV-2 S protein which is predicted by UniProt (UniProtKB P0DTC2) to have a mass of 64.5 kDa (unglycosylated) with additional mass arising from extensive glycosylation [32]. Alternatively, this peak may arise from a fragment of IgA. As such, this signal cannot be unequivocally identified at this time. It should be noted that, in the mass spectrum of viral isolates from cell culture (positive control), an intense peak from bovine serum albumin occurs at 66,600 m/z; this unfortunately would suppress the signal from S2 (if present) into the baseline for the control samples of SARS-CoV-2 (Figure 4).

### 3.4 Estimating LoD on MALDI-ToF

To estimate the sensitivity of the MALDI-ToF protocol, we tested a saliva sample that contained a very low SARS-CoV-2 viral load by quantitative RT-qPCR as described previously [22]. The Ct value of this sample was 36.09 and as such the viral load was less than the quantifiable limit of the assay (600 copies/ml). An aliquot of this saliva sample was processed using the protocol for saliva samples in this study (Section 2.3) and the MALDI-ToF mass spectrum was acquired using the same parameters as gargle samples. The observed signal in the mass spectrum for the potential biomarker peak found between 78,600 to 80,500 m/z was approximately 3 times the baseline noise level (S/N = 3), a commonly accepted value for finding the limit of detection in MALDI-ToF methods (Figure S2). Based on the S/N ratio, we interpreted this sample as being positive for SARS-CoV-2. This result suggests that the MALDI-ToF protocol may indeed be as sensitive as RT-qPCR to detect SARS-CoV-2 in specimens that contain very low viral loads. More studies are necessary to determine the exact limit of detection by the MALDI-ToF protocol.

### 3.5. MALDI-ToF criteria for diagnosis

In order to compare the results of the nasopharyngeal RT-qPCR samples and the protein profiles collected with gargle samples via MALDI-ToF, the AUC was calculated under the seven peak ranges of interest listed in Table 1 for each sample. As stated above, the peak located at 11,150 m/z is most likely to be associated with cystatin A, a resident protein of saliva [19]. The presence of this peak thus was used as an internal control. Table S1 includes the AUC of this peak along with the six other features for all specimens, each titled as the reported RT-qPCR result on NP swabs.

This table (Table S1) was used to sort the samples by AUC values for a given feature from largest to smallest. Sorting in this way showed a reasonable separation between COVID-19 positive and COVID-19 negative specimens for five out of the seven features, including the peaks of S1, S2/immune protein, immunoglobulin heavy/amylase, immunoglobulin heavy doubly-charged, and the biomarker near 112,000 m/z as seen in Figure 5.

**Figure 5.**
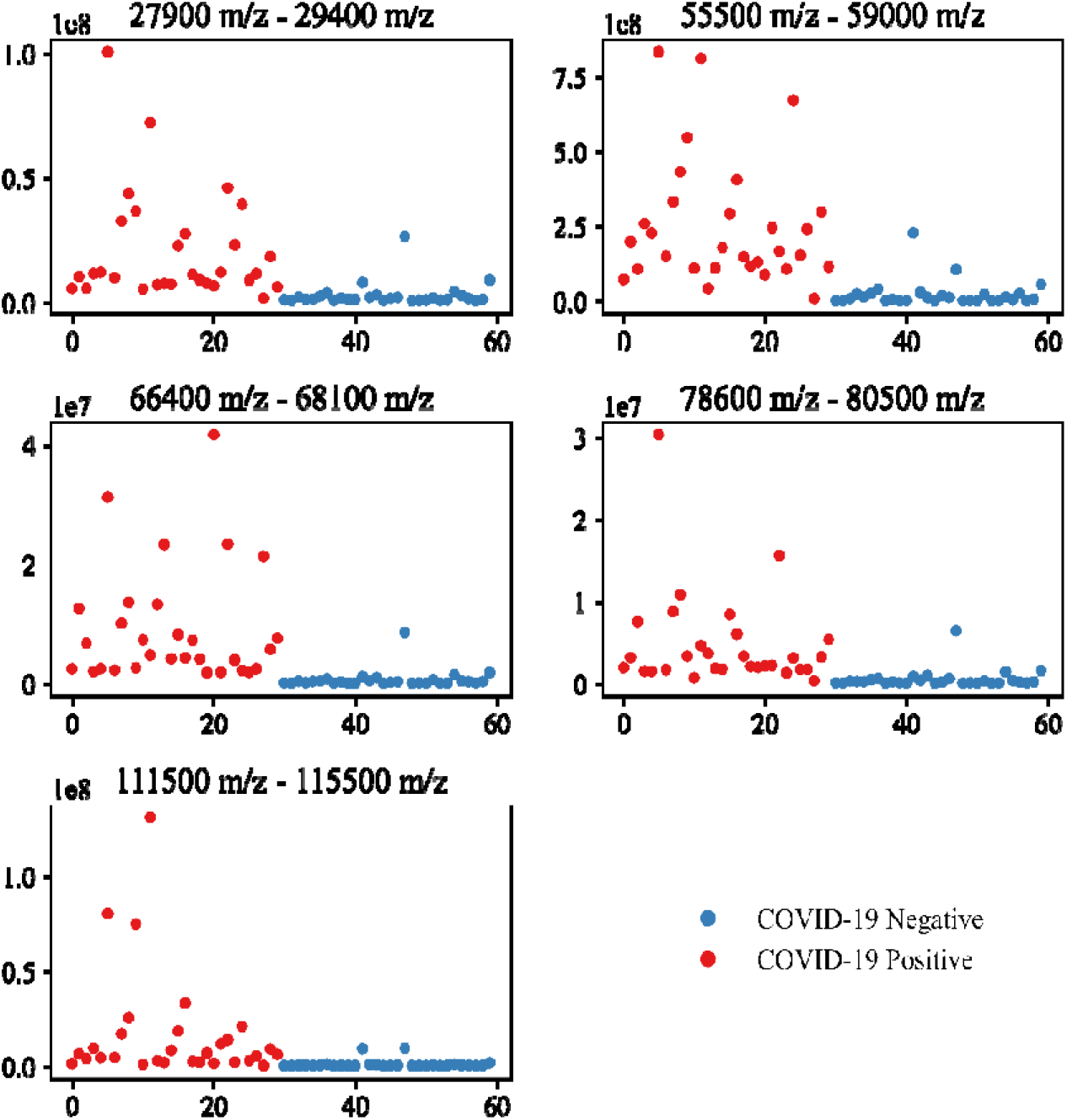
displays AUC values for the 60 gargle samples for the five potential biomarker peak range. The sample data files were labeled 1-60 and the AUC for the biomarker range for each file is depicted. The COVID-19 positive files (red) were clustered and labelled as 1-30 while the COVID-19 negative files (blue) were clustered and labelled as 31-60. A marked difference fo the AUC in the biomarker ranges of COVID-19 positive versus COVID-19 negative samples can be observed.

In order to establish a cutoff threshold for each potential biomarker as it compares to the COVID-19 status, the AUC values for each range were sorted from high to low, as described above, and compared to the disease status. Cutoffs were made that yield the best separation of negative/positive samples. The samples with AUC values above the threshold were assigned MALDI-ToF positive and values below were assigned negative. The cutoff values for a given feature chosen under this criterion are reported in Table 2, along with the percent agreement between RT-qPCR results and our analysis.

**Table 2:**
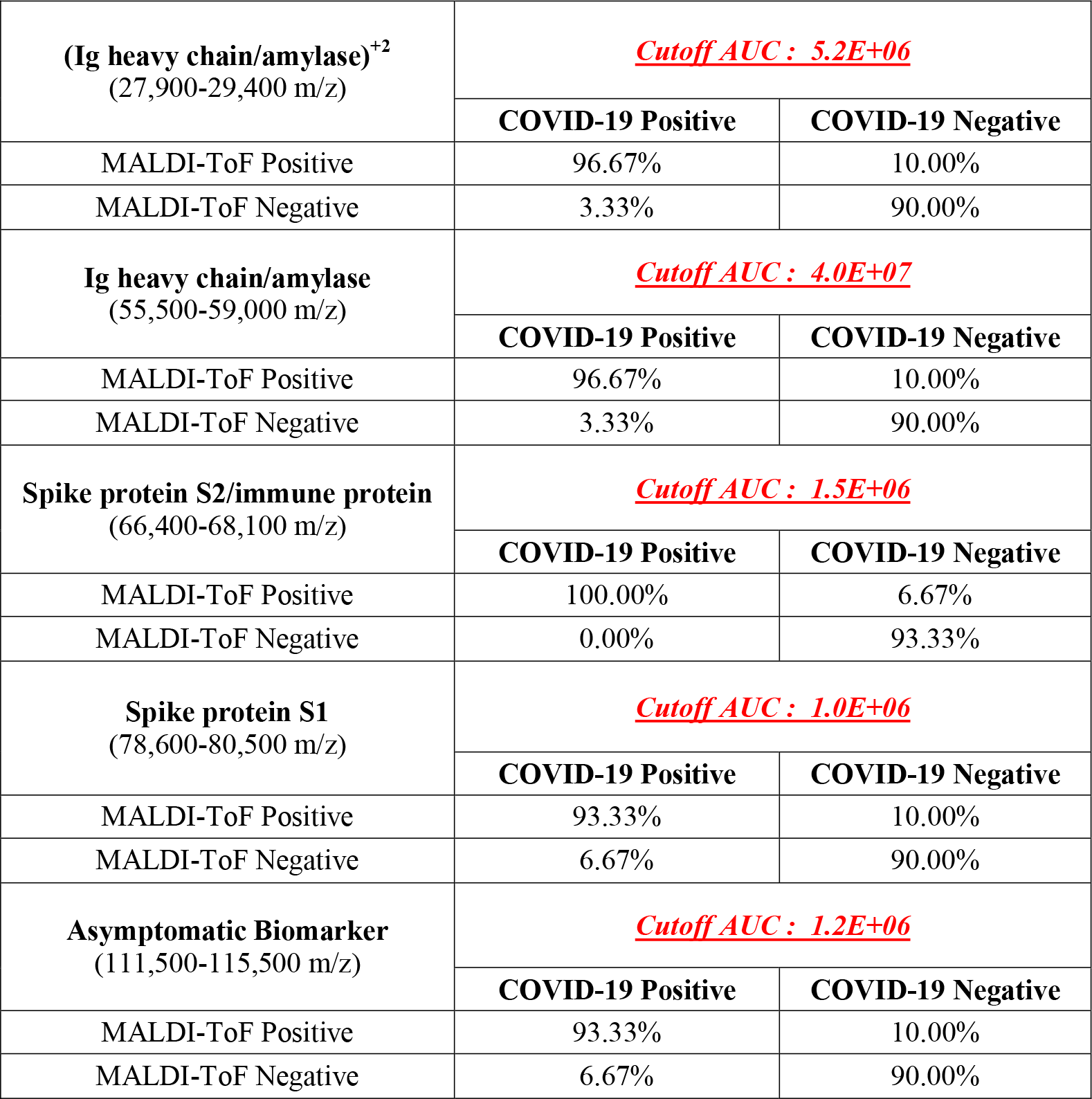
Percent agreement between RT- PCR results and MALDI-ToF results as determined by cutoff AUC values. Cutoff values for each biomarker peak were based on sorting all specimen AUC values for each feature (m/z peak ranges) and setting a threshold AUC value that separated COVID-19 positive from COVID-19 negative results.

For all five potential biomarkers mentioned in Table 2, we achieved 90% and higher agreement with the RT-qPCR results. Although the identities of the proteins are yet to be confirmed, these m/z ranges certainly exhibit a relationship to the COVID-19 status. We refined these observations further with ROC (Receiver Operating characteristic) curve analysis, a commonly used technique in clinical studies to determine sensitivities and specificities at different threshold values of an analyte [33]. The true positive rate (sensitivity) on the Y-axis was plotted against the false positive rate (100-specificity) on the X-axis at each of the observed AUCs for every peak, shown in Figure 6. The analysis clearly demonstrates the ability of the MALDI-ToF assay to discriminate between positive/negative COVID-19 status while providing a quantitative optimization of sensitivity/specificity for each potential biomarker, as summarized in Table 3.

**Table 3:** Summary of ROC curve analysis for each potential biomarker as calculated for AUC of peaks within associated m/z ranges. A peak area threshold is listed for each potential biomarker which optimizes sensitivity and specificity for the MALDI-ToF protocol when compared to COVID-19 status.

| **Potential biomarker**<br>**m/z range** | **Peak area**<br>**threshold** | **AUC-**<br>**ROC** | **Sensitivity**<br>**(%)** | **Specificity**<br>**(%)** |
| --- | --- | --- | --- | --- |
| **Gargle marker**<br>11,140 – 11,160 | >5789021.2 | 0.548 | 40 | 76 |
| **Ig light chains**<br>23,500 – 23,800 | >4629948.3 | 0.756 | 70 | 80 |
| **Ig Heavy**<br>**chain/Amylase<sup>+2</sup>**<br>27,900 – 29,400 | >4368750.1 | 0.941 | 96.67 | 90 |
| **Ig Heavy chain/Amylase**<br>55,500 – 59,000 | >38684701.8 | 0.961 | 96.67 | 90 |
| **Spike protein S2**<br>66,400 – 68,100 | >1429133.2 | 0.976 | 100 | 93.33 |
| **Spike protein S1**<br>78,600 – 80,500 | >937903.9 | 0.954 | 93.33 | 90 |
| **Asymptomatic biomarker**<br>111,500 – 115,500 | >1104743.1 | 0.933 | 93.33 | 90 |

**Figure 6.**
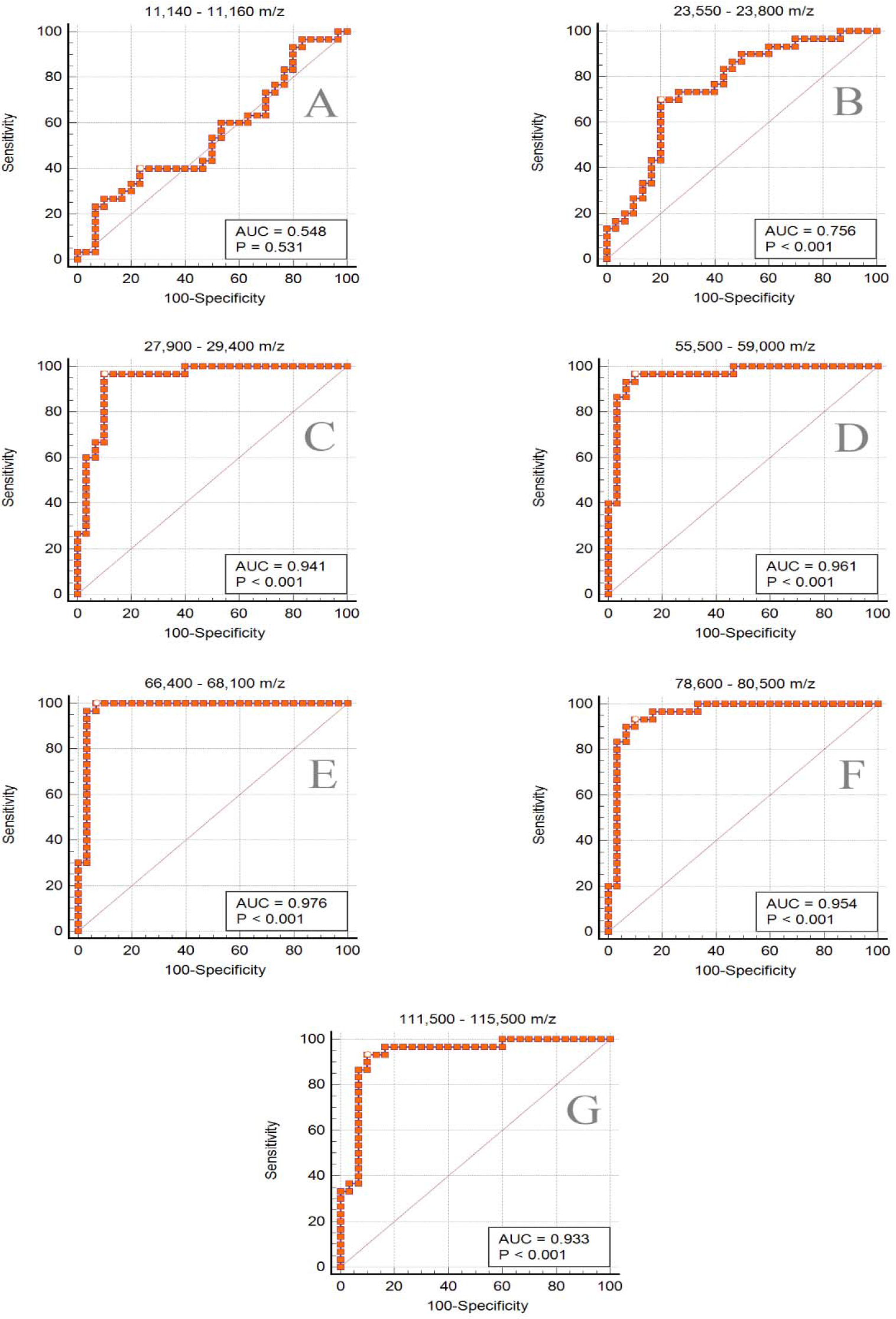
displays the results of ROC curve analysis on the potential protein biomarker range identified by MALDI-ToF from gargle samples. Panel **A-G** present the graphs of true versus false positive rate for each of the seven labeled m/z ranges. In any given panel, each point represents a MALDI-ToF specificity/sensitivity value at a cutoff AUC for each potential biomarker range, with the highest specificity/sensitivity and discriminating power having values near the top left of each plot.

Figure 6 presents the ROC plots of true versus false positive rate for each of the seven m/z ranges listed in Table 1. Panel 6A plots data for the internal quality control biomarker tentatively assigned as cystatin A. The data points lie closely along the diagonal line with the AUC indicating no discrimination between COVID-19 negative and positive samples, as predicted. Panels 6C-G utilize data for the potential biomarkers S1, S2/immune protein, immunoglobulin heavy/amylase, immunoglobulin heavy doubly-charged, and the potential biomarker near 112,000 m/z. These five biomarkers show highly sensitive and specific discrimination between the COVID-19 positive and negative individuals with AUCs ≥ 0.933 and sensitivities and specificities of 93.33-100% and 90-93.33% respectively (Table 3). The closer the AUC is to 1 for ROC curve analysis, the better the model for predicting the disease state. More detailed results of the ROC curve analysis for each potential biomarker can be found in the Supplementary Information. Panel 6B displays the graph for the immunoglobulin light chains m/z region that shows a much weaker discrimination ability; this phenomenon was also observed anecdotally from visual examination of the mass spectra. In summary, the ROC analysis indicates that five peaks in the MALDI-ToF spectrum are capable of highly sensitive and specific discrimination of COVID-19 status in individuals.

It is interesting to note that 89% of the positive samples were from donors who were asymptomatic. This is not surprising given that the student athletes in this study are generally young, healthy individuals. This cohort has been an elusive group to track, particularly early in the pandemic when only symptomatic persons were eligible for RT-qPCR testing. The COVID-19 positive, asymptomatic group were reported to be as contagious as symptomatic persons [34] towards the beginning of the pandemic and had been suggested to be disproportionately responsible for spreading the virus due to a lack of symptoms. This could be of concern in a setting such as a university where communal housing, dining and recreational facilities are predominant. Recent reports, however, cast doubt about a linear relationship between viral loads, symptoms and transmissibility, i.e., how much of an RT-qPCR detected viral load is replicating virus that can be transmitted to other persons is unclear at the present time. Transmissibility will have to be confirmed by viral cultures [5], [35].

## 4. Conclusion

A highly sensitive and specific saliva/gargle test was developed for diagnosing COVID-19 infection using MALDI-ToF mass spectrometry. The method described in this study is relatively rapid, inexpensive and is sensitive enough to detect SARS-CoV-2 infection in samples with very low viral loads. It has advantages over other tests such as non-invasive sampling and the ability to observe both viral proteins and host response. Comparison of COVID-19 status (assessed by RT-qPCR of NP swabs) with MALDI-ToF analysis over a wide range of 2,000 to 200,000 m/z identified five potential biomarkers of COVID-19 infection. These results need to be validated on a larger cohort of samples. It is also crucial to further identify the potential biomarker peaks to understand the relationship of the proteins with the course of the disease. Preliminary studies to identify the peaks were performed by examining human immunoglobulins and heat inactivated SARS-CoV-2 virus by this reported assay and comparing peak masses of these controls to those observed for gargle samples. Further confirmatory analysis such as sequencing of proteins that occur in a healthy and SARS-CoV-2-infected saliva proteome is required. The verification of the identity of the human immune biomarkers may serve as a useful tool for monitoring the immune response under various conditions and stressors. While discrimination of COVID-19 status was based on the AUC of the separate potential biomarkers, we aim to develop machine learning models for achieving unbiased, higher accuracies using multiple peaks in tandem for clinical diagnosis.

## Data Availability

Data will be made available as supplemental material with the peer-reviewed and accepted manuscript or upon request

## 5. Acknowledgements

The authors gratefully acknowledge instrumentation support from Shimadzu Scientific Instruments, Inc. We thank Dr. Ray Iles, MAPSciences, for the kind gift of LBSD-X buffer and Northern Illinois University Athletics (Yordon Center) for assisting us with the sample collection.

The positive control reagent (Figure 4) was deposited by the Centers for Disease Control and Prevention and obtained through BEI Resources, NIAID, NIH: SARS-Related Coronavirus 2, Isolate USA-WA1/2020, Heat Inactivated, NR-52286.

## IRB information

The protocol “Use of MALDI TOF mass spectrometry to analyze SARS-CoV-2 viral proteins” was approved by the Northern Illinois University Institutional Review Board on August 12, 2020.

The protocol “Methods to detect SARS-CoV-2 in saliva without nucleic acid extraction” was approved by Institutional Review Board of the University of Illinois at Chicago on February 11, 2021.

## Abbreviations

ACE2: angiotensin-converting enzyme 2
AUC: area under the curve
COVID-19: coronavirus disease 2019
Ct: cycle threshold
DTT: dithiothreitol
E Protein: envelope protein
EUA: emergency use authorization
FDA: food and drug administration
IgA: immunoglobulin A
IgG: immunoglobulin G
IgM: immunoglobulin M
LoD: limit of detection
LC-MS: liquid chromatography mass spectrometry
M Protein: membrane protein
MALDI-ToF MS: matrix-assisted laser desorption/ionization-time of flight mass spectrometry
N Protein: nucleocapsid protein
NP: nasopharyngeal
RBD: receptor binding domain
RNA: ribonucleic acid
ROC: receiver operating characteristic
RT-qPCR: reverse transcriptase quantitative polymerase chain reaction
S Protein: spike protein
SARS-CoV-2: severe acute respiratory syndrome coronavirus 2
VEP: viral envelope protein
WHO: world health organization

