## Supplementary Information for "MALDI-ToF Protein Profiling as Potential Rapid Diagnostic Platform for COVID-19"

^#^ co-first authors

* corresponding author

Figure S1 ………………………………………………………………………………………………… S2

Figure S2 …………………………………………………………………………………………….…... S2

ROC Curve Results for Peak 11,140 - 11,160 m/z ……………………………………………………… S3

ROC Curve Results for Peak 23,550 - 23,800 m/z ……………………………………………………… S8

ROC Curve Results for Peak 27,900 - 29,400 m/z …………………………………………………..… S13

ROC Curve Results for Peak 55,500 - 59,000 m/z …………………………………………………..… S18

ROC Curve Results for Peak 66,400 - 68,100 m/z …………………………………………………..… S23

ROC Curve Results for Peak 78,600 - 80,500 m/z …………………………………………………..… S28

ROC Curve Results for Peak 111,500-115,500 m/z ………………………………………………...…. S33

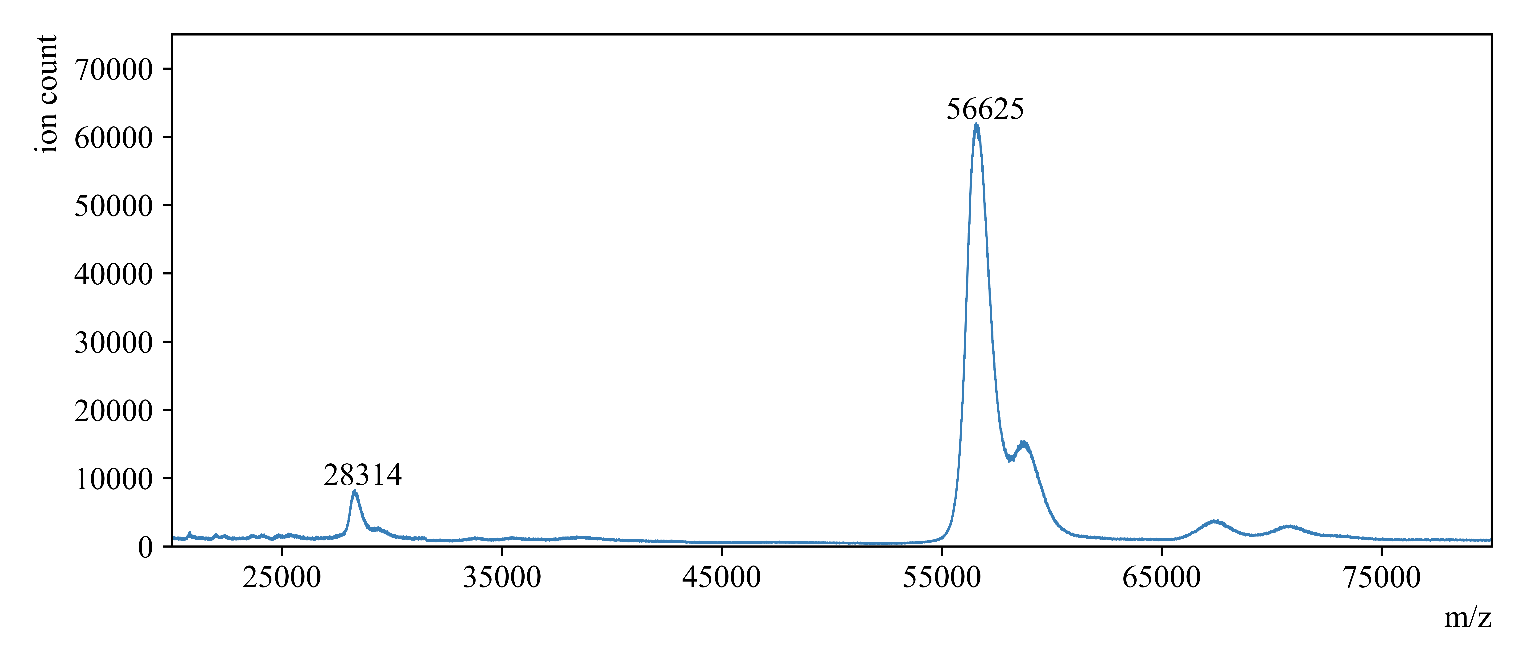

***Figure S1*** *displays the MALDI mass spectrum of human salivary α-amylase, prepared by spotting 100 pmol in LCMS-H_2_O sandwich style as described in main text (Section 2.3).*

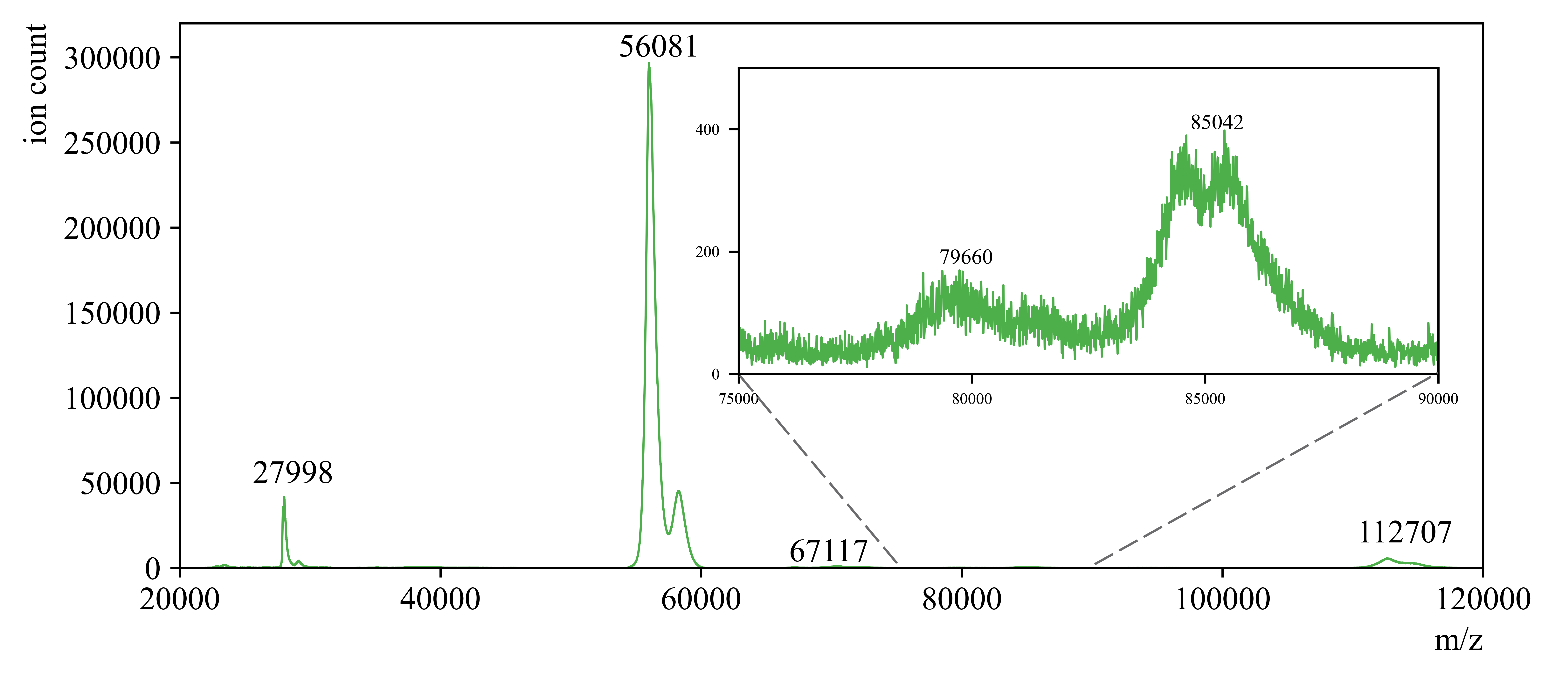

***Figure S2*** *displays the MALDI mass spectrum of a COVID-19 positive saliva sample prepared as in main text (Section 2.3) beyond the limit of quantitation for RT-qPCR [22, main text]. The inset shows the potential biomarker peak in the range of 78,600 – 80,500 m/z having a signal approximately 3 times higher than the baseline (S/N = 3), which is commonly accepted for finding the limit of detection in MALDI-ToF methods.*

ROC curve

| Variable | **11,140-11,160 m/z** |
| --- | --- |
| Classification variable | COVID-19_Status |

| Sample size | 60 |
| --- | --- |
| Positive group ^a^ | 30 (50.00%) |
| Negative group ^b^ | 30 (50.00%) |

^a^ COVID-19 Status = 1
^b^ COVID-19 Status = 0

| Disease prevalence (%) | 10 |
| --- | --- |

#### Area under the ROC curve (AUC)

| Area under the ROC curve (AUC) | 0.548 |
| --- | --- |
| Standard Error ^a^ | 0.0762 |
| 95% Confidence interval ^b^ | 0.414 to 0.677 |
| z statistic | 0.627 |
| Significance level P (Area=0.5) | 0.5306 |

^a^ DeLong et al., 1988

^b^ Binomial exact

#### Youden index

| Youden index J | 0.1667 |
| --- | --- |
| Associated criterion | >5789021.17 |
| Sensitivity | 40.00 |
| Specificity | 76.67 |

#### Criterion values and coordinates of the ROC curve [[Show]](javascript:showdiv('d36','d37','table1');)

#### Criterion values and coordinates of the ROC curve

| Criterion | Sensitivity | 95% CI | Specificity | 95% CI | +LR | 95% CI | -LR | 95% CI | +PV | -PV |
| --- | --- | --- | --- | --- | --- | --- | --- | --- | --- | --- |
| ≥159890.88 | 100.00 | 88.4 - 100.0 | 0.00 | 0.0 - 11.6 | 1.00 | 1.0 - 1.0 |  |  | 10.0 |  |
| >159890.88 | 100.00 | 88.4 - 100.0 | 3.33 | 0.08 - 17.2 | 1.03 | 1.0 - 1.1 | 0.00 |  | 10.3 | 100.0 |
| >322723.81 | 96.67 | 82.8 - 99.9 | 3.33 | 0.08 - 17.2 | 1.00 | 0.9 - 1.1 | 1.00 | 0.07 - 15.3 | 10.0 | 90.0 |
| >362936.63 | 96.67 | 82.8 - 99.9 | 6.67 | 0.8 - 22.1 | 1.04 | 0.9 - 1.2 | 0.50 | 0.05 - 5.2 | 10.3 | 94.7 |
| >442014.2 | 96.67 | 82.8 - 99.9 | 10.00 | 2.1 - 26.5 | 1.07 | 0.9 - 1.2 | 0.33 | 0.04 - 3.0 | 10.7 | 96.4 |
| >524447.88 | 96.67 | 82.8 - 99.9 | 13.33 | 3.8 - 30.7 | 1.12 | 1.0 - 1.3 | 0.25 | 0.03 - 2.1 | 11.0 | 97.3 |
| >557295.19 | 96.67 | 82.8 - 99.9 | 16.67 | 5.6 - 34.7 | 1.16 | 1.0 - 1.4 | 0.20 | 0.02 - 1.6 | 11.4 | 97.8 |
| >671809.88 | 93.33 | 77.9 - 99.2 | 16.67 | 5.6 - 34.7 | 1.12 | 0.9 - 1.3 | 0.40 | 0.08 - 1.9 | 11.1 | 95.7 |
| >673072.04 | 93.33 | 77.9 - 99.2 | 20.00 | 7.7 - 38.6 | 1.17 | 1.0 - 1.4 | 0.33 | 0.07 - 1.5 | 11.5 | 96.4 |
| >749334.7 | 90.00 | 73.5 - 97.9 | 20.00 | 7.7 - 38.6 | 1.12 | 0.9 - 1.4 | 0.50 | 0.1 - 1.8 | 11.1 | 94.7 |
| >878223.15 | 86.67 | 69.3 - 96.2 | 20.00 | 7.7 - 38.6 | 1.08 | 0.9 - 1.4 | 0.67 | 0.2 - 2.1 | 10.7 | 93.1 |
| >994667.79 | 83.33 | 65.3 - 94.4 | 20.00 | 7.7 - 38.6 | 1.04 | 0.8 - 1.3 | 0.83 | 0.3 - 2.4 | 10.4 | 91.5 |
| >1078655.12 | 83.33 | 65.3 - 94.4 | 23.33 | 9.9 - 42.3 | 1.09 | 0.8 - 1.4 | 0.71 | 0.3 - 2.0 | 10.8 | 92.6 |
| >1134277.33 | 80.00 | 61.4 - 92.3 | 23.33 | 9.9 - 42.3 | 1.04 | 0.8 - 1.4 | 0.86 | 0.3 - 2.3 | 10.4 | 91.3 |
| >1341107.49 | 76.67 | 57.7 - 90.1 | 23.33 | 9.9 - 42.3 | 1.00 | 0.8 - 1.3 | 1.00 | 0.4 - 2.5 | 10.0 | 90.0 |
| >1512676.12 | 76.67 | 57.7 - 90.1 | 26.67 | 12.3 - 45.9 | 1.05 | 0.8 - 1.4 | 0.87 | 0.4 - 2.1 | 10.4 | 91.1 |
| >1547428.89 | 73.33 | 54.1 - 87.7 | 26.67 | 12.3 - 45.9 | 1.00 | 0.7 - 1.4 | 1.00 | 0.4 - 2.3 | 10.0 | 90.0 |
| >1558869.43 | 73.33 | 54.1 - 87.7 | 30.00 | 14.7 - 49.4 | 1.05 | 0.8 - 1.4 | 0.89 | 0.4 - 2.0 | 10.4 | 91.0 |
| >1587251.62 | 70.00 | 50.6 - 85.3 | 30.00 | 14.7 - 49.4 | 1.00 | 0.7 - 1.4 | 1.00 | 0.5 - 2.2 | 10.0 | 90.0 |
| >1764022.67 | 66.67 | 47.2 - 82.7 | 30.00 | 14.7 - 49.4 | 0.95 | 0.7 - 1.3 | 1.11 | 0.5 - 2.3 | 9.6 | 89.0 |
| >1801034 | 63.33 | 43.9 - 80.1 | 30.00 | 14.7 - 49.4 | 0.90 | 0.6 - 1.3 | 1.22 | 0.6 - 2.5 | 9.1 | 88.0 |
| >1845886.16 | 63.33 | 43.9 - 80.1 | 33.33 | 17.3 - 52.8 | 0.95 | 0.7 - 1.4 | 1.10 | 0.6 - 2.2 | 9.5 | 89.1 |
| >2188618.88 | 63.33 | 43.9 - 80.1 | 36.67 | 19.9 - 56.1 | 1.00 | 0.7 - 1.5 | 1.00 | 0.5 - 1.9 | 10.0 | 90.0 |
| >2285839.2 | 60.00 | 40.6 - 77.3 | 36.67 | 19.9 - 56.1 | 0.95 | 0.6 - 1.4 | 1.09 | 0.6 - 2.1 | 9.5 | 89.2 |
| >2347210.76 | 60.00 | 40.6 - 77.3 | 40.00 | 22.7 - 59.4 | 1.00 | 0.7 - 1.5 | 1.00 | 0.5 - 1.9 | 10.0 | 90.0 |
| >2474084.34 | 60.00 | 40.6 - 77.3 | 43.33 | 25.5 - 62.6 | 1.06 | 0.7 - 1.6 | 0.92 | 0.5 - 1.7 | 10.5 | 90.7 |
| >3312065.22 | 60.00 | 40.6 - 77.3 | 46.67 | 28.3 - 65.7 | 1.12 | 0.7 - 1.8 | 0.86 | 0.5 - 1.5 | 11.1 | 91.3 |
| >3585193.56999999 | 56.67 | 37.4 - 74.5 | 46.67 | 28.3 - 65.7 | 1.06 | 0.7 - 1.7 | 0.93 | 0.5 - 1.6 | 10.6 | 90.6 |
| >3870139.04 | 53.33 | 34.3 - 71.7 | 46.67 | 28.3 - 65.7 | 1.00 | 0.6 - 1.6 | 1.00 | 0.6 - 1.7 | 10.0 | 90.0 |
| >3975166.79 | 53.33 | 34.3 - 71.7 | 50.00 | 31.3 - 68.7 | 1.07 | 0.7 - 1.7 | 0.93 | 0.6 - 1.6 | 10.6 | 90.6 |
| >4397952.71 | 50.00 | 31.3 - 68.7 | 50.00 | 31.3 - 68.7 | 1.00 | 0.6 - 1.7 | 1.00 | 0.6 - 1.7 | 10.0 | 90.0 |
| >4536305.39 | 46.67 | 28.3 - 65.7 | 50.00 | 31.3 - 68.7 | 0.93 | 0.6 - 1.6 | 1.07 | 0.7 - 1.7 | 9.4 | 89.4 |
| >4646817.9 | 43.33 | 25.5 - 62.6 | 50.00 | 31.3 - 68.7 | 0.87 | 0.5 - 1.5 | 1.13 | 0.7 - 1.8 | 8.8 | 88.8 |
| >4669323.94 | 43.33 | 25.5 - 62.6 | 53.33 | 34.3 - 71.7 | 0.93 | 0.5 - 1.6 | 1.06 | 0.7 - 1.7 | 9.4 | 89.4 |
| >4864463.19 | 40.00 | 22.7 - 59.4 | 53.33 | 34.3 - 71.7 | 0.86 | 0.5 - 1.5 | 1.12 | 0.7 - 1.8 | 8.7 | 88.9 |
| >4982812.85 | 40.00 | 22.7 - 59.4 | 56.67 | 37.4 - 74.5 | 0.92 | 0.5 - 1.7 | 1.06 | 0.7 - 1.6 | 9.3 | 89.5 |
| >5050718.92 | 40.00 | 22.7 - 59.4 | 60.00 | 40.6 - 77.3 | 1.00 | 0.5 - 1.9 | 1.00 | 0.7 - 1.5 | 10.0 | 90.0 |
| >5219483.56 | 40.00 | 22.7 - 59.4 | 63.33 | 43.9 - 80.1 | 1.09 | 0.6 - 2.1 | 0.95 | 0.6 - 1.4 | 10.8 | 90.5 |
| >5378517.79999999 | 40.00 | 22.7 - 59.4 | 66.67 | 47.2 - 82.7 | 1.20 | 0.6 - 2.3 | 0.90 | 0.6 - 1.3 | 11.8 | 90.9 |
| >5536858.6 | 40.00 | 22.7 - 59.4 | 70.00 | 50.6 - 85.3 | 1.33 | 0.7 - 2.7 | 0.86 | 0.6 - 1.2 | 12.9 | 91.3 |
| >5736935.9 | 40.00 | 22.7 - 59.4 | 73.33 | 54.1 - 87.7 | 1.50 | 0.7 - 3.1 | 0.82 | 0.6 - 1.2 | 14.3 | 91.7 |
| >5789021.17 | 40.00 | 22.7 - 59.4 | 76.67 | 57.7 - 90.1 | 1.71 | 0.8 - 3.8 | 0.78 | 0.6 - 1.1 | 16.0 | 92.0 |
| >6352725.84 | 36.67 | 19.9 - 56.1 | 76.67 | 57.7 - 90.1 | 1.57 | 0.7 - 3.5 | 0.83 | 0.6 - 1.2 | 14.9 | 91.6 |
| >6403440.13 | 33.33 | 17.3 - 52.8 | 76.67 | 57.7 - 90.1 | 1.43 | 0.6 - 3.3 | 0.87 | 0.6 - 1.2 | 13.7 | 91.2 |
| >6466488.6 | 33.33 | 17.3 - 52.8 | 80.00 | 61.4 - 92.3 | 1.67 | 0.7 - 4.0 | 0.83 | 0.6 - 1.1 | 15.6 | 91.5 |
| >6774062.12 | 30.00 | 14.7 - 49.4 | 80.00 | 61.4 - 92.3 | 1.50 | 0.6 - 3.7 | 0.88 | 0.7 - 1.2 | 14.3 | 91.1 |
| >6981173.61 | 30.00 | 14.7 - 49.4 | 83.33 | 65.3 - 94.4 | 1.80 | 0.7 - 4.7 | 0.84 | 0.6 - 1.1 | 16.7 | 91.5 |
| >8311104.17 | 26.67 | 12.3 - 45.9 | 83.33 | 65.3 - 94.4 | 1.60 | 0.6 - 4.3 | 0.88 | 0.7 - 1.2 | 15.1 | 91.1 |
| >8581229.15 | 26.67 | 12.3 - 45.9 | 86.67 | 69.3 - 96.2 | 2.00 | 0.7 - 5.9 | 0.85 | 0.7 - 1.1 | 18.2 | 91.4 |
| >9076136.99 | 26.67 | 12.3 - 45.9 | 90.00 | 73.5 - 97.9 | 2.67 | 0.8 - 9.1 | 0.81 | 0.6 - 1.0 | 22.9 | 91.7 |
| >9250253.18999999 | 23.33 | 9.9 - 42.3 | 90.00 | 73.5 - 97.9 | 2.33 | 0.7 - 8.2 | 0.85 | 0.7 - 1.1 | 20.6 | 91.4 |
| >9590278.40999999 | 23.33 | 9.9 - 42.3 | 93.33 | 77.9 - 99.2 | 3.50 | 0.8 - 15.5 | 0.82 | 0.7 - 1.0 | 28.0 | 91.6 |
| >10572057.3999999 | 20.00 | 7.7 - 38.6 | 93.33 | 77.9 - 99.2 | 3.00 | 0.7 - 13.7 | 0.86 | 0.7 - 1.0 | 25.0 | 91.3 |
| >11084113.4 | 16.67 | 5.6 - 34.7 | 93.33 | 77.9 - 99.2 | 2.50 | 0.5 - 11.9 | 0.89 | 0.7 - 1.1 | 21.7 | 91.0 |
| >11831319.46 | 13.33 | 3.8 - 30.7 | 93.33 | 77.9 - 99.2 | 2.00 | 0.4 - 10.1 | 0.93 | 0.8 - 1.1 | 18.2 | 90.6 |
| >14291115.77 | 10.00 | 2.1 - 26.5 | 93.33 | 77.9 - 99.2 | 1.50 | 0.3 - 8.3 | 0.96 | 0.8 - 1.1 | 14.3 | 90.3 |
| >15625591.75 | 6.67 | 0.8 - 22.1 | 93.33 | 77.9 - 99.2 | 1.00 | 0.2 - 6.6 | 1.00 | 0.9 - 1.1 | 10.0 | 90.0 |
| >17167011.18 | 3.33 | 0.08 - 17.2 | 93.33 | 77.9 - 99.2 | 0.50 | 0.05 - 5.2 | 1.04 | 0.9 - 1.2 | 5.3 | 89.7 |
| >18215945.6 | 3.33 | 0.08 - 17.2 | 96.67 | 82.8 - 99.9 | 1.00 | 0.07 - 15.3 | 1.00 | 0.9 - 1.1 | 10.0 | 90.0 |
| >20344828.1 | 3.33 | 0.08 - 17.2 | 100.00 | 88.4 - 100.0 |  |  | 0.97 | 0.9 - 1.0 | 100.0 | 90.3 |
| >26869593.72 | 0.00 | 0.0 - 11.6 | 100.00 | 88.4 - 100.0 |  |  | 1.00 | 1.0 - 1.0 |  | 90.0 |

### ROC curve

| Variable | **23,550-23,800 m/z** |
| --- | --- |
| Classification variable | COVID-19_Status |

| Sample size | 60 |
| --- | --- |
| Positive group ^a^ | 30 (50.00%) |
| Negative group ^b^ | 30 (50.00%) |

^a^ COVID-19_Status = 1
^b^ COVID-19_Status = 0

| Disease prevalence (%) | 10 |
| --- | --- |

#### Area under the ROC curve (AUC)

| Area under the ROC curve (AUC) | 0.756 |
| --- | --- |
| Standard Error ^a^ | 0.0639 |
| 95% Confidence interval ^b^ | 0.627 to 0.857 |
| z statistic | 3.997 |
| Significance level P (Area=0.5) | 0.0001 |

^a^ DeLong et al., 1988

^b^ Binomial exact

#### Youden index

| Youden index J | 0.5000 |
| --- | --- |
| Associated criterion | >4629948.31 |
| Sensitivity | 70.00 |
| Specificity | 80.00 |

#### Criterion values and coordinates of the ROC curve [[Show]](javascript:showdiv('d24','d25','table1');)

#### Criterion values and coordinates of the ROC curve

| Criterion | Sensitivity | 95% CI | Specificity | 95% CI | +LR | 95% CI | -LR | 95% CI | +PV | -PV |
| --- | --- | --- | --- | --- | --- | --- | --- | --- | --- | --- |
| ≥159449.62 | 100.00 | 88.4 - 100.0 | 0.00 | 0.0 - 11.6 | 1.00 | 1.0 - 1.0 |  |  | 10.0 |  |
| >159449.62 | 100.00 | 88.4 - 100.0 | 3.33 | 0.08 - 17.2 | 1.03 | 1.0 - 1.1 | 0.00 |  | 10.3 | 100.0 |
| >453339.8 | 100.00 | 88.4 - 100.0 | 6.67 | 0.8 - 22.1 | 1.07 | 1.0 - 1.2 | 0.00 |  | 10.6 | 100.0 |
| >585464.81 | 100.00 | 88.4 - 100.0 | 10.00 | 2.1 - 26.5 | 1.11 | 1.0 - 1.3 | 0.00 |  | 11.0 | 100.0 |
| >702489.61 | 100.00 | 88.4 - 100.0 | 13.33 | 3.8 - 30.7 | 1.15 | 1.0 - 1.3 | 0.00 |  | 11.4 | 100.0 |
| >706498.42 | 96.67 | 82.8 - 99.9 | 13.33 | 3.8 - 30.7 | 1.12 | 1.0 - 1.3 | 0.25 | 0.03 - 2.1 | 11.0 | 97.3 |
| >801219.29 | 96.67 | 82.8 - 99.9 | 16.67 | 5.6 - 34.7 | 1.16 | 1.0 - 1.4 | 0.20 | 0.02 - 1.6 | 11.4 | 97.8 |
| >802902.79 | 96.67 | 82.8 - 99.9 | 20.00 | 7.7 - 38.6 | 1.21 | 1.0 - 1.5 | 0.17 | 0.02 - 1.3 | 11.8 | 98.2 |
| >1104210.57 | 96.67 | 82.8 - 99.9 | 23.33 | 9.9 - 42.3 | 1.26 | 1.0 - 1.6 | 0.14 | 0.02 - 1.1 | 12.3 | 98.4 |
| >1314531.56 | 96.67 | 82.8 - 99.9 | 26.67 | 12.3 - 45.9 | 1.32 | 1.1 - 1.7 | 0.13 | 0.02 - 0.9 | 12.8 | 98.6 |
| >1458385.27 | 96.67 | 82.8 - 99.9 | 30.00 | 14.7 - 49.4 | 1.38 | 1.1 - 1.8 | 0.11 | 0.01 - 0.8 | 13.3 | 98.8 |
| >1561772.7 | 93.33 | 77.9 - 99.2 | 30.00 | 14.7 - 49.4 | 1.33 | 1.0 - 1.7 | 0.22 | 0.05 - 0.9 | 12.9 | 97.6 |
| >1608486.26 | 93.33 | 77.9 - 99.2 | 33.33 | 17.3 - 52.8 | 1.40 | 1.1 - 1.8 | 0.20 | 0.05 - 0.8 | 13.5 | 97.8 |
| >2037132.28 | 93.33 | 77.9 - 99.2 | 36.67 | 19.9 - 56.1 | 1.47 | 1.1 - 2.0 | 0.18 | 0.04 - 0.8 | 14.1 | 98.0 |
| >2121087.11 | 93.33 | 77.9 - 99.2 | 40.00 | 22.7 - 59.4 | 1.56 | 1.1 - 2.1 | 0.17 | 0.04 - 0.7 | 14.7 | 98.2 |
| >2189089.04 | 90.00 | 73.5 - 97.9 | 40.00 | 22.7 - 59.4 | 1.50 | 1.1 - 2.1 | 0.25 | 0.08 - 0.8 | 14.3 | 97.3 |
| >2216325.04 | 90.00 | 73.5 - 97.9 | 43.33 | 25.5 - 62.6 | 1.59 | 1.1 - 2.2 | 0.23 | 0.07 - 0.7 | 15.0 | 97.5 |
| >2253400.28 | 90.00 | 73.5 - 97.9 | 46.67 | 28.3 - 65.7 | 1.69 | 1.2 - 2.4 | 0.21 | 0.07 - 0.7 | 15.8 | 97.7 |
| >2259279.15 | 90.00 | 73.5 - 97.9 | 50.00 | 31.3 - 68.7 | 1.80 | 1.2 - 2.6 | 0.20 | 0.06 - 0.6 | 16.7 | 97.8 |
| >2393189.75 | 86.67 | 69.3 - 96.2 | 50.00 | 31.3 - 68.7 | 1.73 | 1.2 - 2.5 | 0.27 | 0.1 - 0.7 | 16.1 | 97.1 |
| >2595949.98 | 86.67 | 69.3 - 96.2 | 53.33 | 34.3 - 71.7 | 1.86 | 1.2 - 2.8 | 0.25 | 0.09 - 0.7 | 17.1 | 97.3 |
| >2694751.01 | 83.33 | 65.3 - 94.4 | 53.33 | 34.3 - 71.7 | 1.79 | 1.2 - 2.7 | 0.31 | 0.1 - 0.7 | 16.6 | 96.6 |
| >2822991.06 | 83.33 | 65.3 - 94.4 | 56.67 | 37.4 - 74.5 | 1.92 | 1.2 - 3.0 | 0.29 | 0.1 - 0.7 | 17.6 | 96.8 |
| >2968551.32 | 80.00 | 61.4 - 92.3 | 56.67 | 37.4 - 74.5 | 1.85 | 1.2 - 2.9 | 0.35 | 0.2 - 0.8 | 17.0 | 96.2 |
| >3014662.86 | 76.67 | 57.7 - 90.1 | 56.67 | 37.4 - 74.5 | 1.77 | 1.1 - 2.8 | 0.41 | 0.2 - 0.8 | 16.4 | 95.6 |
| >3076083.84999999 | 76.67 | 57.7 - 90.1 | 60.00 | 40.6 - 77.3 | 1.92 | 1.2 - 3.1 | 0.39 | 0.2 - 0.8 | 17.6 | 95.9 |
| >3246717.32 | 73.33 | 54.1 - 87.7 | 60.00 | 40.6 - 77.3 | 1.83 | 1.1 - 3.0 | 0.44 | 0.2 - 0.9 | 16.9 | 95.3 |
| >3434927.52 | 73.33 | 54.1 - 87.7 | 63.33 | 43.9 - 80.1 | 2.00 | 1.2 - 3.4 | 0.42 | 0.2 - 0.8 | 18.2 | 95.5 |
| >3912357.69 | 73.33 | 54.1 - 87.7 | 66.67 | 47.2 - 82.7 | 2.20 | 1.3 - 3.8 | 0.40 | 0.2 - 0.8 | 19.6 | 95.7 |
| >3928660.38 | 73.33 | 54.1 - 87.7 | 70.00 | 50.6 - 85.3 | 2.44 | 1.4 - 4.4 | 0.38 | 0.2 - 0.7 | 21.4 | 95.9 |
| >4045446.58 | 73.33 | 54.1 - 87.7 | 73.33 | 54.1 - 87.7 | 2.75 | 1.5 - 5.2 | 0.36 | 0.2 - 0.7 | 23.4 | 96.1 |
| >4172259.75 | 70.00 | 50.6 - 85.3 | 73.33 | 54.1 - 87.7 | 2.62 | 1.4 - 5.0 | 0.41 | 0.2 - 0.7 | 22.6 | 95.7 |
| >4324914.63 | 70.00 | 50.6 - 85.3 | 76.67 | 57.7 - 90.1 | 3.00 | 1.5 - 6.0 | 0.39 | 0.2 - 0.7 | 25.0 | 95.8 |
| >4629948.31 | 70.00 | 50.6 - 85.3 | 80.00 | 61.4 - 92.3 | 3.50 | 1.6 - 7.4 | 0.38 | 0.2 - 0.7 | 28.0 | 96.0 |
| >4687543.99 | 66.67 | 47.2 - 82.7 | 80.00 | 61.4 - 92.3 | 3.33 | 1.6 - 7.1 | 0.42 | 0.2 - 0.7 | 27.0 | 95.6 |
| >4818675.87 | 63.33 | 43.9 - 80.1 | 80.00 | 61.4 - 92.3 | 3.17 | 1.5 - 6.8 | 0.46 | 0.3 - 0.8 | 26.0 | 95.2 |
| >5186073.38999999 | 60.00 | 40.6 - 77.3 | 80.00 | 61.4 - 92.3 | 3.00 | 1.4 - 6.5 | 0.50 | 0.3 - 0.8 | 25.0 | 94.7 |
| >5211306.97 | 56.67 | 37.4 - 74.5 | 80.00 | 61.4 - 92.3 | 2.83 | 1.3 - 6.2 | 0.54 | 0.3 - 0.8 | 23.9 | 94.3 |
| >5933539.31 | 53.33 | 34.3 - 71.7 | 80.00 | 61.4 - 92.3 | 2.67 | 1.2 - 5.9 | 0.58 | 0.4 - 0.9 | 22.9 | 93.9 |
| >6178337.53 | 50.00 | 31.3 - 68.7 | 80.00 | 61.4 - 92.3 | 2.50 | 1.1 - 5.6 | 0.63 | 0.4 - 0.9 | 21.7 | 93.5 |
| >6797262.47 | 46.67 | 28.3 - 65.7 | 80.00 | 61.4 - 92.3 | 2.33 | 1.0 - 5.3 | 0.67 | 0.5 - 1.0 | 20.6 | 93.1 |
| >7678774.65 | 43.33 | 25.5 - 62.6 | 80.00 | 61.4 - 92.3 | 2.17 | 1.0 - 4.9 | 0.71 | 0.5 - 1.0 | 19.4 | 92.7 |
| >9298196.46 | 43.33 | 25.5 - 62.6 | 83.33 | 65.3 - 94.4 | 2.60 | 1.1 - 6.4 | 0.68 | 0.5 - 1.0 | 22.4 | 93.0 |
| >9756892.89 | 40.00 | 22.7 - 59.4 | 83.33 | 65.3 - 94.4 | 2.40 | 1.0 - 6.0 | 0.72 | 0.5 - 1.0 | 21.1 | 92.6 |
| >10202152.8599999 | 36.67 | 19.9 - 56.1 | 83.33 | 65.3 - 94.4 | 2.20 | 0.9 - 5.6 | 0.76 | 0.6 - 1.0 | 19.6 | 92.2 |
| >10504907.58 | 33.33 | 17.3 - 52.8 | 83.33 | 65.3 - 94.4 | 2.00 | 0.8 - 5.2 | 0.80 | 0.6 - 1.1 | 18.2 | 91.8 |
| >10624400.51 | 33.33 | 17.3 - 52.8 | 86.67 | 69.3 - 96.2 | 2.50 | 0.9 - 7.1 | 0.77 | 0.6 - 1.0 | 21.7 | 92.1 |
| >12150136.8899999 | 30.00 | 14.7 - 49.4 | 86.67 | 69.3 - 96.2 | 2.25 | 0.8 - 6.5 | 0.81 | 0.6 - 1.1 | 20.0 | 91.8 |
| >12396326.7199999 | 26.67 | 12.3 - 45.9 | 86.67 | 69.3 - 96.2 | 2.00 | 0.7 - 5.9 | 0.85 | 0.7 - 1.1 | 18.2 | 91.4 |
| >12404154.4699999 | 26.67 | 12.3 - 45.9 | 90.00 | 73.5 - 97.9 | 2.67 | 0.8 - 9.1 | 0.81 | 0.6 - 1.0 | 22.9 | 91.7 |
| >13539527.6099999 | 23.33 | 9.9 - 42.3 | 90.00 | 73.5 - 97.9 | 2.33 | 0.7 - 8.2 | 0.85 | 0.7 - 1.1 | 20.6 | 91.4 |
| >13889254.25 | 20.00 | 7.7 - 38.6 | 90.00 | 73.5 - 97.9 | 2.00 | 0.6 - 7.3 | 0.89 | 0.7 - 1.1 | 18.2 | 91.0 |
| >15416438.0999999 | 20.00 | 7.7 - 38.6 | 93.33 | 77.9 - 99.2 | 3.00 | 0.7 - 13.7 | 0.86 | 0.7 - 1.0 | 25.0 | 91.3 |
| >17273876.2799999 | 16.67 | 5.6 - 34.7 | 93.33 | 77.9 - 99.2 | 2.50 | 0.5 - 11.9 | 0.89 | 0.7 - 1.1 | 21.7 | 91.0 |
| >20429145.4799999 | 16.67 | 5.6 - 34.7 | 96.67 | 82.8 - 99.9 | 5.00 | 0.6 - 40.3 | 0.86 | 0.7 - 1.0 | 35.7 | 91.3 |
| >21944752.6999999 | 13.33 | 3.8 - 30.7 | 96.67 | 82.8 - 99.9 | 4.00 | 0.5 - 33.7 | 0.90 | 0.8 - 1.0 | 30.8 | 90.9 |
| >23458395.2799999 | 13.33 | 3.8 - 30.7 | 100.00 | 88.4 - 100.0 |  |  | 0.87 | 0.8 - 1.0 | 100.0 | 91.2 |
| >30153135.5799999 | 10.00 | 2.1 - 26.5 | 100.00 | 88.4 - 100.0 |  |  | 0.90 | 0.8 - 1.0 | 100.0 | 90.9 |
| >32768452.39 | 6.67 | 0.8 - 22.1 | 100.00 | 88.4 - 100.0 |  |  | 0.93 | 0.8 - 1.0 | 100.0 | 90.6 |
| >41113644.23 | 3.33 | 0.08 - 17.2 | 100.00 | 88.4 - 100.0 |  |  | 0.97 | 0.9 - 1.0 | 100.0 | 90.3 |
| >42649240.47 | 0.00 | 0.0 - 11.6 | 100.00 | 88.4 - 100.0 |  |  | 1.00 | 1.0 - 1.0 |  | 90.0 |

### ROC curve

| Variable | **27,900-29,400 m/z** |
| --- | --- |
| Classification variable | COVID-19_status |

| Sample size | 60 |
| --- | --- |
| Positive group ^a^ | 30 (50.00%) |
| Negative group ^b^ | 30 (50.00%) |

^a^ COVID-19_status = 1
^b^ COVID-19_status = 0

| Disease prevalence (%) | 10 |
| --- | --- |

#### Area under the ROC curve (AUC)

| Area under the ROC curve (AUC) | 0.941 |
| --- | --- |
| Standard Error ^a^ | 0.0318 |
| 95% Confidence interval ^b^ | 0.849 to 0.985 |
| z statistic | 13.863 |
| Significance level P (Area=0.5) | <0.0001 |

^a^ DeLong et al., 1988

^b^ Binomial exact

#### Youden index

| Youden index J | 0.8667 |
| --- | --- |
| Associated criterion | >4368750.14 |
| Sensitivity | 96.67 |
| Specificity | 90.00 |

#### Criterion values and coordinates of the ROC curve [[Show]](javascript:showdiv('d26','d27','table1');)

#### Criterion values and coordinates of the ROC curve

| Criterion | Sensitivity | 95% CI | Specificity | 95% CI | +LR | 95% CI | -LR | 95% CI | +PV | -PV |
| --- | --- | --- | --- | --- | --- | --- | --- | --- | --- | --- |
| ≥630931.52 | 100.00 | 88.4 - 100.0 | 0.00 | 0.0 - 11.6 | 1.00 | 1.0 - 1.0 |  |  | 10.0 |  |
| >630931.52 | 100.00 | 88.4 - 100.0 | 3.33 | 0.08 - 17.2 | 1.03 | 1.0 - 1.1 | 0.00 |  | 10.3 | 100.0 |
| >639059.67 | 100.00 | 88.4 - 100.0 | 6.67 | 0.8 - 22.1 | 1.07 | 1.0 - 1.2 | 0.00 |  | 10.6 | 100.0 |
| >718581.59 | 100.00 | 88.4 - 100.0 | 10.00 | 2.1 - 26.5 | 1.11 | 1.0 - 1.3 | 0.00 |  | 11.0 | 100.0 |
| >730035.19 | 100.00 | 88.4 - 100.0 | 13.33 | 3.8 - 30.7 | 1.15 | 1.0 - 1.3 | 0.00 |  | 11.4 | 100.0 |
| >750752.33 | 100.00 | 88.4 - 100.0 | 16.67 | 5.6 - 34.7 | 1.20 | 1.0 - 1.4 | 0.00 |  | 11.8 | 100.0 |
| >762900.94 | 100.00 | 88.4 - 100.0 | 20.00 | 7.7 - 38.6 | 1.25 | 1.0 - 1.5 | 0.00 |  | 12.2 | 100.0 |
| >763103.95 | 100.00 | 88.4 - 100.0 | 23.33 | 9.9 - 42.3 | 1.30 | 1.1 - 1.6 | 0.00 |  | 12.7 | 100.0 |
| >817983.08 | 100.00 | 88.4 - 100.0 | 26.67 | 12.3 - 45.9 | 1.36 | 1.1 - 1.7 | 0.00 |  | 13.2 | 100.0 |
| >912210.04 | 100.00 | 88.4 - 100.0 | 30.00 | 14.7 - 49.4 | 1.43 | 1.1 - 1.8 | 0.00 |  | 13.7 | 100.0 |
| >947559.05 | 100.00 | 88.4 - 100.0 | 33.33 | 17.3 - 52.8 | 1.50 | 1.2 - 1.9 | 0.00 |  | 14.3 | 100.0 |
| >998537.63 | 100.00 | 88.4 - 100.0 | 36.67 | 19.9 - 56.1 | 1.58 | 1.2 - 2.1 | 0.00 |  | 14.9 | 100.0 |
| >1020287.05 | 100.00 | 88.4 - 100.0 | 40.00 | 22.7 - 59.4 | 1.67 | 1.2 - 2.2 | 0.00 |  | 15.6 | 100.0 |
| >1047720.3 | 100.00 | 88.4 - 100.0 | 43.33 | 25.5 - 62.6 | 1.76 | 1.3 - 2.4 | 0.00 |  | 16.4 | 100.0 |
| >1097378.35 | 100.00 | 88.4 - 100.0 | 46.67 | 28.3 - 65.7 | 1.87 | 1.3 - 2.6 | 0.00 |  | 17.2 | 100.0 |
| >1117511.73 | 100.00 | 88.4 - 100.0 | 50.00 | 31.3 - 68.7 | 2.00 | 1.4 - 2.9 | 0.00 |  | 18.2 | 100.0 |
| >1327742.06 | 100.00 | 88.4 - 100.0 | 53.33 | 34.3 - 71.7 | 2.14 | 1.5 - 3.1 | 0.00 |  | 19.2 | 100.0 |
| >1397570.58 | 100.00 | 88.4 - 100.0 | 56.67 | 37.4 - 74.5 | 2.31 | 1.5 - 3.5 | 0.00 |  | 20.4 | 100.0 |
| >1483844.85 | 100.00 | 88.4 - 100.0 | 60.00 | 40.6 - 77.3 | 2.50 | 1.6 - 3.9 | 0.00 |  | 21.7 | 100.0 |
| >1490153.73 | 96.67 | 82.8 - 99.9 | 60.00 | 40.6 - 77.3 | 2.42 | 1.6 - 3.8 | 0.056 | 0.008 - 0.4 | 21.2 | 99.4 |
| >1662883.5 | 96.67 | 82.8 - 99.9 | 63.33 | 43.9 - 80.1 | 2.64 | 1.6 - 4.2 | 0.053 | 0.008 - 0.4 | 22.7 | 99.4 |
| >1781175.65 | 96.67 | 82.8 - 99.9 | 66.67 | 47.2 - 82.7 | 2.90 | 1.7 - 4.8 | 0.050 | 0.007 - 0.3 | 24.4 | 99.4 |
| >1794325.59 | 96.67 | 82.8 - 99.9 | 70.00 | 50.6 - 85.3 | 3.22 | 1.9 - 5.6 | 0.048 | 0.007 - 0.3 | 26.4 | 99.5 |
| >1910774.7 | 96.67 | 82.8 - 99.9 | 73.33 | 54.1 - 87.7 | 3.62 | 2.0 - 6.6 | 0.045 | 0.007 - 0.3 | 28.7 | 99.5 |
| >2279216.79 | 96.67 | 82.8 - 99.9 | 76.67 | 57.7 - 90.1 | 4.14 | 2.2 - 8.0 | 0.043 | 0.006 - 0.3 | 31.5 | 99.5 |
| >2541697.94 | 96.67 | 82.8 - 99.9 | 80.00 | 61.4 - 92.3 | 4.83 | 2.4 - 9.9 | 0.042 | 0.006 - 0.3 | 34.9 | 99.5 |
| >2755349.8 | 96.67 | 82.8 - 99.9 | 83.33 | 65.3 - 94.4 | 5.80 | 2.6 - 12.9 | 0.040 | 0.006 - 0.3 | 39.2 | 99.6 |
| >3592520.48 | 96.67 | 82.8 - 99.9 | 86.67 | 69.3 - 96.2 | 7.25 | 2.9 - 18.1 | 0.038 | 0.006 - 0.3 | 44.6 | 99.6 |
| >4368750.14 | 96.67 | 82.8 - 99.9 | 90.00 | 73.5 - 97.9 | 9.67 | 3.3 - 28.3 | 0.037 | 0.005 - 0.3 | 51.8 | 99.6 |
| >5276438.35 | 93.33 | 77.9 - 99.2 | 90.00 | 73.5 - 97.9 | 9.33 | 3.2 - 27.4 | 0.074 | 0.02 - 0.3 | 50.9 | 99.2 |
| >5513971.31 | 90.00 | 73.5 - 97.9 | 90.00 | 73.5 - 97.9 | 9.00 | 3.1 - 26.5 | 0.11 | 0.04 - 0.3 | 50.0 | 98.8 |
| >5579502.68 | 86.67 | 69.3 - 96.2 | 90.00 | 73.5 - 97.9 | 8.67 | 2.9 - 25.6 | 0.15 | 0.06 - 0.4 | 49.1 | 98.4 |
| >6044535.65 | 83.33 | 65.3 - 94.4 | 90.00 | 73.5 - 97.9 | 8.33 | 2.8 - 24.7 | 0.19 | 0.08 - 0.4 | 48.1 | 98.0 |
| >6486850.93 | 80.00 | 61.4 - 92.3 | 90.00 | 73.5 - 97.9 | 8.00 | 2.7 - 23.8 | 0.22 | 0.1 - 0.5 | 47.1 | 97.6 |
| >6901230.18 | 76.67 | 57.7 - 90.1 | 90.00 | 73.5 - 97.9 | 7.67 | 2.6 - 22.8 | 0.26 | 0.1 - 0.5 | 46.0 | 97.2 |
| >7129267 | 73.33 | 54.1 - 87.7 | 90.00 | 73.5 - 97.9 | 7.33 | 2.5 - 21.9 | 0.30 | 0.2 - 0.5 | 44.9 | 96.8 |
| >7356395.27 | 70.00 | 50.6 - 85.3 | 90.00 | 73.5 - 97.9 | 7.00 | 2.3 - 21.0 | 0.33 | 0.2 - 0.6 | 43.7 | 96.4 |
| >7423702.04 | 66.67 | 47.2 - 82.7 | 90.00 | 73.5 - 97.9 | 6.67 | 2.2 - 20.1 | 0.37 | 0.2 - 0.6 | 42.6 | 96.0 |
| >7794134.99 | 66.67 | 47.2 - 82.7 | 93.33 | 77.9 - 99.2 | 10.00 | 2.6 - 39.1 | 0.36 | 0.2 - 0.6 | 52.6 | 96.2 |
| >8674898.70999999 | 63.33 | 43.9 - 80.1 | 93.33 | 77.9 - 99.2 | 9.50 | 2.4 - 37.2 | 0.39 | 0.2 - 0.6 | 51.4 | 95.8 |
| >8795181.72999999 | 60.00 | 40.6 - 77.3 | 93.33 | 77.9 - 99.2 | 9.00 | 2.3 - 35.4 | 0.43 | 0.3 - 0.7 | 50.0 | 95.5 |
| >8820209.91999999 | 60.00 | 40.6 - 77.3 | 96.67 | 82.8 - 99.9 | 18.00 | 2.6 - 126.4 | 0.41 | 0.3 - 0.6 | 66.7 | 95.6 |
| >9840930.9 | 56.67 | 37.4 - 74.5 | 96.67 | 82.8 - 99.9 | 17.00 | 2.4 - 119.8 | 0.45 | 0.3 - 0.7 | 65.4 | 95.3 |
| >10223012.58 | 53.33 | 34.3 - 71.7 | 96.67 | 82.8 - 99.9 | 16.00 | 2.3 - 113.1 | 0.48 | 0.3 - 0.7 | 64.0 | 94.9 |
| >11109324.2699999 | 50.00 | 31.3 - 68.7 | 96.67 | 82.8 - 99.9 | 15.00 | 2.1 - 106.5 | 0.52 | 0.4 - 0.7 | 62.5 | 94.6 |
| >11444807.1299999 | 46.67 | 28.3 - 65.7 | 96.67 | 82.8 - 99.9 | 14.00 | 2.0 - 99.9 | 0.55 | 0.4 - 0.8 | 60.9 | 94.2 |
| >11468929.6299999 | 43.33 | 25.5 - 62.6 | 96.67 | 82.8 - 99.9 | 13.00 | 1.8 - 93.2 | 0.59 | 0.4 - 0.8 | 59.1 | 93.9 |
| >11930600.38 | 40.00 | 22.7 - 59.4 | 96.67 | 82.8 - 99.9 | 12.00 | 1.7 - 86.6 | 0.62 | 0.5 - 0.8 | 57.1 | 93.5 |
| >11963137.86 | 36.67 | 19.9 - 56.1 | 96.67 | 82.8 - 99.9 | 11.00 | 1.5 - 80.0 | 0.66 | 0.5 - 0.9 | 55.0 | 93.2 |
| >18220235.75 | 33.33 | 17.3 - 52.8 | 96.67 | 82.8 - 99.9 | 10.00 | 1.4 - 73.3 | 0.69 | 0.5 - 0.9 | 52.6 | 92.9 |
| >22527996.68 | 30.00 | 14.7 - 49.4 | 96.67 | 82.8 - 99.9 | 9.00 | 1.2 - 66.7 | 0.72 | 0.6 - 0.9 | 50.0 | 92.6 |
| >22902477.63 | 26.67 | 12.3 - 45.9 | 96.67 | 82.8 - 99.9 | 8.00 | 1.1 - 60.1 | 0.76 | 0.6 - 1.0 | 47.1 | 92.2 |
| >26340243.5199999 | 26.67 | 12.3 - 45.9 | 100.00 | 88.4 - 100.0 |  |  | 0.73 | 0.6 - 0.9 | 100.0 | 92.5 |
| >27374821.9699999 | 23.33 | 9.9 - 42.3 | 100.00 | 88.4 - 100.0 |  |  | 0.77 | 0.6 - 0.9 | 100.0 | 92.2 |
| >32495989.3999999 | 20.00 | 7.7 - 38.6 | 100.00 | 88.4 - 100.0 |  |  | 0.80 | 0.7 - 1.0 | 100.0 | 91.8 |
| >36470803.8499999 | 16.67 | 5.6 - 34.7 | 100.00 | 88.4 - 100.0 |  |  | 0.83 | 0.7 - 1.0 | 100.0 | 91.5 |
| >39140624.27 | 13.33 | 3.8 - 30.7 | 100.00 | 88.4 - 100.0 |  |  | 0.87 | 0.8 - 1.0 | 100.0 | 91.2 |
| >43431862.59 | 10.00 | 2.1 - 26.5 | 100.00 | 88.4 - 100.0 |  |  | 0.90 | 0.8 - 1.0 | 100.0 | 90.9 |
| >45904716.17 | 6.67 | 0.8 - 22.1 | 100.00 | 88.4 - 100.0 |  |  | 0.93 | 0.8 - 1.0 | 100.0 | 90.6 |
| >72120088.29 | 3.33 | 0.08 - 17.2 | 100.00 | 88.4 - 100.0 |  |  | 0.97 | 0.9 - 1.0 | 100.0 | 90.3 |
| >100430198.53 | 0.00 | 0.0 - 11.6 | 100.00 | 88.4 - 100.0 |  |  | 1.00 | 1.0 - 1.0 |  | 90.0 |

### ROC curve

| Variable | **55,500-59,000 m/z** |
| --- | --- |
| Classification variable | COVID-19_Status |

| Sample size | 60 |
| --- | --- |
| Positive group ^a^ | 30 (50.00%) |
| Negative group ^b^ | 30 (50.00%) |

^a^ COVID-19_Status = 1
^b^ COVID-19_Status = 0

| Disease prevalence (%) | 10 |
| --- | --- |

#### Area under the ROC curve (AUC)

| Area under the ROC curve (AUC) | 0.961 |
| --- | --- |
| Standard Error ^a^ | 0.0253 |
| 95% Confidence interval ^b^ | 0.877 to 0.994 |
| z statistic | 18.241 |
| Significance level P (Area=0.5) | <0.0001 |

^a^ DeLong et al., 1988

^b^ Binomial exact

#### Youden index

| Youden index J | 0.8667 |
| --- | --- |
| Associated criterion | >38684701.83 |
| Sensitivity | 96.67 |
| Specificity | 90.00 |

#### Criterion values and coordinates of the ROC curve [[Show]](javascript:showdiv('d28','d29','table1');)

#### Criterion values and coordinates of the ROC curve [[Hide]](javascript:hidediv('d28','d29','table1');)

| Criterion | Sensitivity | 95% CI | Specificity | 95% CI | +LR | 95% CI | -LR | 95% CI | +PV | -PV |
| --- | --- | --- | --- | --- | --- | --- | --- | --- | --- | --- |
| ≥256954.4 | 100.00 | 88.4 - 100.0 | 0.00 | 0.0 - 11.6 | 1.00 | 1.0 - 1.0 |  |  | 10.0 |  |
| >256954.4 | 100.00 | 88.4 - 100.0 | 3.33 | 0.08 - 17.2 | 1.03 | 1.0 - 1.1 | 0.00 |  | 10.3 | 100.0 |
| >297808.95 | 100.00 | 88.4 - 100.0 | 6.67 | 0.8 - 22.1 | 1.07 | 1.0 - 1.2 | 0.00 |  | 10.6 | 100.0 |
| >308806.3 | 100.00 | 88.4 - 100.0 | 10.00 | 2.1 - 26.5 | 1.11 | 1.0 - 1.3 | 0.00 |  | 11.0 | 100.0 |
| >366142.09 | 100.00 | 88.4 - 100.0 | 13.33 | 3.8 - 30.7 | 1.15 | 1.0 - 1.3 | 0.00 |  | 11.4 | 100.0 |
| >471439.85 | 100.00 | 88.4 - 100.0 | 16.67 | 5.6 - 34.7 | 1.20 | 1.0 - 1.4 | 0.00 |  | 11.8 | 100.0 |
| >539909.4 | 100.00 | 88.4 - 100.0 | 20.00 | 7.7 - 38.6 | 1.25 | 1.0 - 1.5 | 0.00 |  | 12.2 | 100.0 |
| >550303.27 | 100.00 | 88.4 - 100.0 | 23.33 | 9.9 - 42.3 | 1.30 | 1.1 - 1.6 | 0.00 |  | 12.7 | 100.0 |
| >553470.41 | 100.00 | 88.4 - 100.0 | 26.67 | 12.3 - 45.9 | 1.36 | 1.1 - 1.7 | 0.00 |  | 13.2 | 100.0 |
| >670580.67 | 100.00 | 88.4 - 100.0 | 30.00 | 14.7 - 49.4 | 1.43 | 1.1 - 1.8 | 0.00 |  | 13.7 | 100.0 |
| >766360.15 | 100.00 | 88.4 - 100.0 | 33.33 | 17.3 - 52.8 | 1.50 | 1.2 - 1.9 | 0.00 |  | 14.3 | 100.0 |
| >790223.63 | 100.00 | 88.4 - 100.0 | 36.67 | 19.9 - 56.1 | 1.58 | 1.2 - 2.1 | 0.00 |  | 14.9 | 100.0 |
| >793424.23 | 100.00 | 88.4 - 100.0 | 40.00 | 22.7 - 59.4 | 1.67 | 1.2 - 2.2 | 0.00 |  | 15.6 | 100.0 |
| >2559522.41999999 | 100.00 | 88.4 - 100.0 | 43.33 | 25.5 - 62.6 | 1.76 | 1.3 - 2.4 | 0.00 |  | 16.4 | 100.0 |
| >3736161.21 | 100.00 | 88.4 - 100.0 | 46.67 | 28.3 - 65.7 | 1.87 | 1.3 - 2.6 | 0.00 |  | 17.2 | 100.0 |
| >4446438.42 | 100.00 | 88.4 - 100.0 | 50.00 | 31.3 - 68.7 | 2.00 | 1.4 - 2.9 | 0.00 |  | 18.2 | 100.0 |
| >5712739.10999999 | 100.00 | 88.4 - 100.0 | 53.33 | 34.3 - 71.7 | 2.14 | 1.5 - 3.1 | 0.00 |  | 19.2 | 100.0 |
| >6173447.33 | 96.67 | 82.8 - 99.9 | 53.33 | 34.3 - 71.7 | 2.07 | 1.4 - 3.1 | 0.063 | 0.009 - 0.4 | 18.7 | 99.3 |
| >9542646.59 | 96.67 | 82.8 - 99.9 | 56.67 | 37.4 - 74.5 | 2.23 | 1.5 - 3.4 | 0.059 | 0.008 - 0.4 | 19.9 | 99.4 |
| >9970136.26999999 | 96.67 | 82.8 - 99.9 | 60.00 | 40.6 - 77.3 | 2.42 | 1.6 - 3.8 | 0.056 | 0.008 - 0.4 | 21.2 | 99.4 |
| >11615255.46 | 96.67 | 82.8 - 99.9 | 63.33 | 43.9 - 80.1 | 2.64 | 1.6 - 4.2 | 0.053 | 0.008 - 0.4 | 22.7 | 99.4 |
| >11874407.0299999 | 96.67 | 82.8 - 99.9 | 66.67 | 47.2 - 82.7 | 2.90 | 1.7 - 4.8 | 0.050 | 0.007 - 0.3 | 24.4 | 99.4 |
| >16254814.2099999 | 96.67 | 82.8 - 99.9 | 70.00 | 50.6 - 85.3 | 3.22 | 1.9 - 5.6 | 0.048 | 0.007 - 0.3 | 26.4 | 99.5 |
| >20221196.9399999 | 96.67 | 82.8 - 99.9 | 73.33 | 54.1 - 87.7 | 3.62 | 2.0 - 6.6 | 0.045 | 0.007 - 0.3 | 28.7 | 99.5 |
| >21072339.01 | 96.67 | 82.8 - 99.9 | 76.67 | 57.7 - 90.1 | 4.14 | 2.2 - 8.0 | 0.043 | 0.006 - 0.3 | 31.5 | 99.5 |
| >22719992.4799999 | 96.67 | 82.8 - 99.9 | 80.00 | 61.4 - 92.3 | 4.83 | 2.4 - 9.9 | 0.042 | 0.006 - 0.3 | 34.9 | 99.5 |
| >22798234.17 | 96.67 | 82.8 - 99.9 | 83.33 | 65.3 - 94.4 | 5.80 | 2.6 - 12.9 | 0.040 | 0.006 - 0.3 | 39.2 | 99.6 |
| >27738066.8699999 | 96.67 | 82.8 - 99.9 | 86.67 | 69.3 - 96.2 | 7.25 | 2.9 - 18.1 | 0.038 | 0.006 - 0.3 | 44.6 | 99.6 |
| >38684701.83 | 96.67 | 82.8 - 99.9 | 90.00 | 73.5 - 97.9 | 9.67 | 3.3 - 28.3 | 0.037 | 0.005 - 0.3 | 51.8 | 99.6 |
| >40504446.23 | 93.33 | 77.9 - 99.2 | 90.00 | 73.5 - 97.9 | 9.33 | 3.2 - 27.4 | 0.074 | 0.02 - 0.3 | 50.9 | 99.2 |
| >53418070.64 | 93.33 | 77.9 - 99.2 | 93.33 | 77.9 - 99.2 | 14.00 | 3.7 - 53.6 | 0.071 | 0.02 - 0.3 | 60.9 | 99.2 |
| >71107336.02 | 90.00 | 73.5 - 97.9 | 93.33 | 77.9 - 99.2 | 13.50 | 3.5 - 51.8 | 0.11 | 0.04 - 0.3 | 60.0 | 98.8 |
| >86702576.6299999 | 86.67 | 69.3 - 96.2 | 93.33 | 77.9 - 99.2 | 13.00 | 3.4 - 50.0 | 0.14 | 0.06 - 0.4 | 59.1 | 98.4 |
| >103506633.55 | 86.67 | 69.3 - 96.2 | 96.67 | 82.8 - 99.9 | 26.00 | 3.8 - 179.5 | 0.14 | 0.06 - 0.3 | 74.3 | 98.5 |
| >105091078.4 | 83.33 | 65.3 - 94.4 | 96.67 | 82.8 - 99.9 | 25.00 | 3.6 - 172.9 | 0.17 | 0.08 - 0.4 | 73.5 | 98.1 |
| >105214790.479999 | 80.00 | 61.4 - 92.3 | 96.67 | 82.8 - 99.9 | 24.00 | 3.5 - 166.2 | 0.21 | 0.1 - 0.4 | 72.7 | 97.8 |
| >108074295.079999 | 76.67 | 57.7 - 90.1 | 96.67 | 82.8 - 99.9 | 23.00 | 3.3 - 159.6 | 0.24 | 0.1 - 0.5 | 71.9 | 97.4 |
| >108486154.15 | 73.33 | 54.1 - 87.7 | 96.67 | 82.8 - 99.9 | 22.00 | 3.2 - 153.0 | 0.28 | 0.2 - 0.5 | 71.0 | 97.0 |
| >112983350.38 | 70.00 | 50.6 - 85.3 | 96.67 | 82.8 - 99.9 | 21.00 | 3.0 - 146.3 | 0.31 | 0.2 - 0.5 | 70.0 | 96.7 |
| >115483568.71 | 66.67 | 47.2 - 82.7 | 96.67 | 82.8 - 99.9 | 20.00 | 2.9 - 139.7 | 0.34 | 0.2 - 0.6 | 69.0 | 96.3 |
| >128986752.469999 | 63.33 | 43.9 - 80.1 | 96.67 | 82.8 - 99.9 | 19.00 | 2.7 - 133.0 | 0.38 | 0.2 - 0.6 | 67.9 | 96.0 |
| >145773903.38 | 60.00 | 40.6 - 77.3 | 96.67 | 82.8 - 99.9 | 18.00 | 2.6 - 126.4 | 0.41 | 0.3 - 0.6 | 66.7 | 95.6 |
| >147747161.009999 | 56.67 | 37.4 - 74.5 | 96.67 | 82.8 - 99.9 | 17.00 | 2.4 - 119.8 | 0.45 | 0.3 - 0.7 | 65.4 | 95.3 |
| >151694114.549999 | 53.33 | 34.3 - 71.7 | 96.67 | 82.8 - 99.9 | 16.00 | 2.3 - 113.1 | 0.48 | 0.3 - 0.7 | 64.0 | 94.9 |
| >165474963.139999 | 50.00 | 31.3 - 68.7 | 96.67 | 82.8 - 99.9 | 15.00 | 2.1 - 106.5 | 0.52 | 0.4 - 0.7 | 62.5 | 94.6 |
| >177300871.119999 | 46.67 | 28.3 - 65.7 | 96.67 | 82.8 - 99.9 | 14.00 | 2.0 - 99.9 | 0.55 | 0.4 - 0.8 | 60.9 | 94.2 |
| >197116625.37 | 43.33 | 25.5 - 62.6 | 96.67 | 82.8 - 99.9 | 13.00 | 1.8 - 93.2 | 0.59 | 0.4 - 0.8 | 59.1 | 93.9 |
| >225105367.179999 | 40.00 | 22.7 - 59.4 | 96.67 | 82.8 - 99.9 | 12.00 | 1.7 - 86.6 | 0.62 | 0.5 - 0.8 | 57.1 | 93.5 |
| >225755976.629999 | 40.00 | 22.7 - 59.4 | 100.00 | 88.4 - 100.0 |  |  | 0.60 | 0.4 - 0.8 | 100.0 | 93.8 |
| >239260135.66 | 36.67 | 19.9 - 56.1 | 100.00 | 88.4 - 100.0 |  |  | 0.63 | 0.5 - 0.8 | 100.0 | 93.4 |
| >243459018.99 | 33.33 | 17.3 - 52.8 | 100.00 | 88.4 - 100.0 |  |  | 0.67 | 0.5 - 0.9 | 100.0 | 93.1 |
| >257222000.219999 | 30.00 | 14.7 - 49.4 | 100.00 | 88.4 - 100.0 |  |  | 0.70 | 0.6 - 0.9 | 100.0 | 92.8 |
| >290918258.889999 | 26.67 | 12.3 - 45.9 | 100.00 | 88.4 - 100.0 |  |  | 0.73 | 0.6 - 0.9 | 100.0 | 92.5 |
| >296960624.74 | 23.33 | 9.9 - 42.3 | 100.00 | 88.4 - 100.0 |  |  | 0.77 | 0.6 - 0.9 | 100.0 | 92.2 |
| >330940678.669999 | 20.00 | 7.7 - 38.6 | 100.00 | 88.4 - 100.0 |  |  | 0.80 | 0.7 - 1.0 | 100.0 | 91.8 |
| >404182887.889999 | 16.67 | 5.6 - 34.7 | 100.00 | 88.4 - 100.0 |  |  | 0.83 | 0.7 - 1.0 | 100.0 | 91.5 |
| >430927844.6 | 13.33 | 3.8 - 30.7 | 100.00 | 88.4 - 100.0 |  |  | 0.87 | 0.8 - 1.0 | 100.0 | 91.2 |
| >545638715.75 | 10.00 | 2.1 - 26.5 | 100.00 | 88.4 - 100.0 |  |  | 0.90 | 0.8 - 1.0 | 100.0 | 90.9 |
| >670876172.02 | 6.67 | 0.8 - 22.1 | 100.00 | 88.4 - 100.0 |  |  | 0.93 | 0.8 - 1.0 | 100.0 | 90.6 |
| >810196903.509999 | 3.33 | 0.08 - 17.2 | 100.00 | 88.4 - 100.0 |  |  | 0.97 | 0.9 - 1.0 | 100.0 | 90.3 |
| >832059261.79 | 0.00 | 0.0 - 11.6 | 100.00 | 88.4 - 100.0 |  |  | 1.00 | 1.0 - 1.0 |  | 90.0 |

### ROC curve

| Variable | **66,400-68,100 m/z** |
| --- | --- |
| Classification variable | COVID-19_Status |

| Sample size | 60 |
| --- | --- |
| Positive group ^a^ | 30 (50.00%) |
| Negative group ^b^ | 30 (50.00%) |

^a^ COVID-19_Status = 1
^b^ COVID-19_Status = 0

| Disease prevalence (%) | 10 |
| --- | --- |

#### Area under the ROC curve (AUC)

| Area under the ROC curve (AUC) | 0.976 |
| --- | --- |
| Standard Error ^a^ | 0.0235 |
| 95% Confidence interval ^b^ | 0.898 to 0.998 |
| z statistic | 20.206 |
| Significance level P (Area=0.5) | <0.0001 |

^a^ DeLong et al., 1988

^b^ Binomial exact

#### Youden index

| Youden index J | 0.9333 |
| --- | --- |
| Associated criterion | >1429133.22 |
| Sensitivity | 100.00 |
| Specificity | 93.33 |

#### Criterion values and coordinates of the ROC curve [[Show]](javascript:showdiv('d30','d31','table1');)

#### Criterion values and coordinates of the ROC curve

| Criterion | Sensitivity | 95% CI | Specificity | 95% CI | +LR | 95% CI | -LR | 95% CI | +PV | -PV |
| --- | --- | --- | --- | --- | --- | --- | --- | --- | --- | --- |
| ≥22561.35 | 100.00 | 88.4 - 100.0 | 0.00 | 0.0 - 11.6 | 1.00 | 1.0 - 1.0 |  |  | 10.0 |  |
| >22561.35 | 100.00 | 88.4 - 100.0 | 3.33 | 0.08 - 17.2 | 1.03 | 1.0 - 1.1 | 0.00 |  | 10.3 | 100.0 |
| >28525.05 | 100.00 | 88.4 - 100.0 | 6.67 | 0.8 - 22.1 | 1.07 | 1.0 - 1.2 | 0.00 |  | 10.6 | 100.0 |
| >37785.99 | 100.00 | 88.4 - 100.0 | 10.00 | 2.1 - 26.5 | 1.11 | 1.0 - 1.3 | 0.00 |  | 11.0 | 100.0 |
| >40299.68 | 100.00 | 88.4 - 100.0 | 13.33 | 3.8 - 30.7 | 1.15 | 1.0 - 1.3 | 0.00 |  | 11.4 | 100.0 |
| >52512.92 | 100.00 | 88.4 - 100.0 | 16.67 | 5.6 - 34.7 | 1.20 | 1.0 - 1.4 | 0.00 |  | 11.8 | 100.0 |
| >58233.52 | 100.00 | 88.4 - 100.0 | 20.00 | 7.7 - 38.6 | 1.25 | 1.0 - 1.5 | 0.00 |  | 12.2 | 100.0 |
| >58272.76 | 100.00 | 88.4 - 100.0 | 23.33 | 9.9 - 42.3 | 1.30 | 1.1 - 1.6 | 0.00 |  | 12.7 | 100.0 |
| >60539.49 | 100.00 | 88.4 - 100.0 | 26.67 | 12.3 - 45.9 | 1.36 | 1.1 - 1.7 | 0.00 |  | 13.2 | 100.0 |
| >83114.18 | 100.00 | 88.4 - 100.0 | 30.00 | 14.7 - 49.4 | 1.43 | 1.1 - 1.8 | 0.00 |  | 13.7 | 100.0 |
| >88943.36 | 100.00 | 88.4 - 100.0 | 33.33 | 17.3 - 52.8 | 1.50 | 1.2 - 1.9 | 0.00 |  | 14.3 | 100.0 |
| >103623.42 | 100.00 | 88.4 - 100.0 | 36.67 | 19.9 - 56.1 | 1.58 | 1.2 - 2.1 | 0.00 |  | 14.9 | 100.0 |
| >116492.66 | 100.00 | 88.4 - 100.0 | 40.00 | 22.7 - 59.4 | 1.67 | 1.2 - 2.2 | 0.00 |  | 15.6 | 100.0 |
| >139118.36 | 100.00 | 88.4 - 100.0 | 43.33 | 25.5 - 62.6 | 1.76 | 1.3 - 2.4 | 0.00 |  | 16.4 | 100.0 |
| >210991.59 | 100.00 | 88.4 - 100.0 | 46.67 | 28.3 - 65.7 | 1.87 | 1.3 - 2.6 | 0.00 |  | 17.2 | 100.0 |
| >277960.37 | 100.00 | 88.4 - 100.0 | 50.00 | 31.3 - 68.7 | 2.00 | 1.4 - 2.9 | 0.00 |  | 18.2 | 100.0 |
| >304653.63 | 100.00 | 88.4 - 100.0 | 53.33 | 34.3 - 71.7 | 2.14 | 1.5 - 3.1 | 0.00 |  | 19.2 | 100.0 |
| >327100.51 | 100.00 | 88.4 - 100.0 | 56.67 | 37.4 - 74.5 | 2.31 | 1.5 - 3.5 | 0.00 |  | 20.4 | 100.0 |
| >329660.97 | 100.00 | 88.4 - 100.0 | 60.00 | 40.6 - 77.3 | 2.50 | 1.6 - 3.9 | 0.00 |  | 21.7 | 100.0 |
| >359786.96 | 100.00 | 88.4 - 100.0 | 63.33 | 43.9 - 80.1 | 2.73 | 1.7 - 4.4 | 0.00 |  | 23.3 | 100.0 |
| >381442.89 | 100.00 | 88.4 - 100.0 | 66.67 | 47.2 - 82.7 | 3.00 | 1.8 - 5.0 | 0.00 |  | 25.0 | 100.0 |
| >403291.93 | 100.00 | 88.4 - 100.0 | 70.00 | 50.6 - 85.3 | 3.33 | 1.9 - 5.8 | 0.00 |  | 27.0 | 100.0 |
| >437193.66 | 100.00 | 88.4 - 100.0 | 73.33 | 54.1 - 87.7 | 3.75 | 2.1 - 6.8 | 0.00 |  | 29.4 | 100.0 |
| >472090.28 | 100.00 | 88.4 - 100.0 | 76.67 | 57.7 - 90.1 | 4.29 | 2.2 - 8.2 | 0.00 |  | 32.3 | 100.0 |
| >597883.53 | 100.00 | 88.4 - 100.0 | 80.00 | 61.4 - 92.3 | 5.00 | 2.4 - 10.2 | 0.00 |  | 35.7 | 100.0 |
| >738373.38 | 100.00 | 88.4 - 100.0 | 83.33 | 65.3 - 94.4 | 6.00 | 2.7 - 13.4 | 0.00 |  | 40.0 | 100.0 |
| >935044.86 | 100.00 | 88.4 - 100.0 | 86.67 | 69.3 - 96.2 | 7.50 | 3.0 - 18.7 | 0.00 |  | 45.5 | 100.0 |
| >1110421.01 | 100.00 | 88.4 - 100.0 | 90.00 | 73.5 - 97.9 | 10.00 | 3.4 - 29.3 | 0.00 |  | 52.6 | 100.0 |
| >1429133.22 | 100.00 | 88.4 - 100.0 | 93.33 | 77.9 - 99.2 | 15.00 | 3.9 - 57.2 | 0.00 |  | 62.5 | 100.0 |
| >1792055.83 | 96.67 | 82.8 - 99.9 | 93.33 | 77.9 - 99.2 | 14.50 | 3.8 - 55.4 | 0.036 | 0.005 - 0.2 | 61.7 | 99.6 |
| >1792743.51 | 96.67 | 82.8 - 99.9 | 96.67 | 82.8 - 99.9 | 29.00 | 4.2 - 199.4 | 0.034 | 0.005 - 0.2 | 76.3 | 99.6 |
| >1846839.78 | 93.33 | 77.9 - 99.2 | 96.67 | 82.8 - 99.9 | 28.00 | 4.1 - 192.8 | 0.069 | 0.02 - 0.3 | 75.7 | 99.2 |
| >1896989.05 | 90.00 | 73.5 - 97.9 | 96.67 | 82.8 - 99.9 | 27.00 | 3.9 - 186.2 | 0.10 | 0.04 - 0.3 | 75.0 | 98.9 |
| >2034540.89 | 86.67 | 69.3 - 96.2 | 96.67 | 82.8 - 99.9 | 26.00 | 3.8 - 179.5 | 0.14 | 0.06 - 0.3 | 74.3 | 98.5 |
| >2178650.49 | 83.33 | 65.3 - 94.4 | 96.67 | 82.8 - 99.9 | 25.00 | 3.6 - 172.9 | 0.17 | 0.08 - 0.4 | 73.5 | 98.1 |
| >2248694.1 | 80.00 | 61.4 - 92.3 | 96.67 | 82.8 - 99.9 | 24.00 | 3.5 - 166.2 | 0.21 | 0.1 - 0.4 | 72.7 | 97.8 |
| >2453125.05 | 76.67 | 57.7 - 90.1 | 96.67 | 82.8 - 99.9 | 23.00 | 3.3 - 159.6 | 0.24 | 0.1 - 0.5 | 71.9 | 97.4 |
| >2466973.65 | 73.33 | 54.1 - 87.7 | 96.67 | 82.8 - 99.9 | 22.00 | 3.2 - 153.0 | 0.28 | 0.2 - 0.5 | 71.0 | 97.0 |
| >2487606.13 | 70.00 | 50.6 - 85.3 | 96.67 | 82.8 - 99.9 | 21.00 | 3.0 - 146.3 | 0.31 | 0.2 - 0.5 | 70.0 | 96.7 |
| >2625038.66999999 | 66.67 | 47.2 - 82.7 | 96.67 | 82.8 - 99.9 | 20.00 | 2.9 - 139.7 | 0.34 | 0.2 - 0.6 | 69.0 | 96.3 |
| >3920308.02999999 | 63.33 | 43.9 - 80.1 | 96.67 | 82.8 - 99.9 | 19.00 | 2.7 - 133.0 | 0.38 | 0.2 - 0.6 | 67.9 | 96.0 |
| >4100731.25 | 60.00 | 40.6 - 77.3 | 96.67 | 82.8 - 99.9 | 18.00 | 2.6 - 126.4 | 0.41 | 0.3 - 0.6 | 66.7 | 95.6 |
| >4160793.31 | 56.67 | 37.4 - 74.5 | 96.67 | 82.8 - 99.9 | 17.00 | 2.4 - 119.8 | 0.45 | 0.3 - 0.7 | 65.4 | 95.3 |
| >4328447.09999999 | 53.33 | 34.3 - 71.7 | 96.67 | 82.8 - 99.9 | 16.00 | 2.3 - 113.1 | 0.48 | 0.3 - 0.7 | 64.0 | 94.9 |
| >4770213.3 | 50.00 | 31.3 - 68.7 | 96.67 | 82.8 - 99.9 | 15.00 | 2.1 - 106.5 | 0.52 | 0.4 - 0.7 | 62.5 | 94.6 |
| >5684587.56 | 46.67 | 28.3 - 65.7 | 96.67 | 82.8 - 99.9 | 14.00 | 2.0 - 99.9 | 0.55 | 0.4 - 0.8 | 60.9 | 94.2 |
| >6767579.83 | 43.33 | 25.5 - 62.6 | 96.67 | 82.8 - 99.9 | 13.00 | 1.8 - 93.2 | 0.59 | 0.4 - 0.8 | 59.1 | 93.9 |
| >7211758.21 | 40.00 | 22.7 - 59.4 | 96.67 | 82.8 - 99.9 | 12.00 | 1.7 - 86.6 | 0.62 | 0.5 - 0.8 | 57.1 | 93.5 |
| >7286941.67 | 36.67 | 19.9 - 56.1 | 96.67 | 82.8 - 99.9 | 11.00 | 1.5 - 80.0 | 0.66 | 0.5 - 0.9 | 55.0 | 93.2 |
| >7540364.35 | 33.33 | 17.3 - 52.8 | 96.67 | 82.8 - 99.9 | 10.00 | 1.4 - 73.3 | 0.69 | 0.5 - 0.9 | 52.6 | 92.9 |
| >8173005.7 | 30.00 | 14.7 - 49.4 | 96.67 | 82.8 - 99.9 | 9.00 | 1.2 - 66.7 | 0.72 | 0.6 - 0.9 | 50.0 | 92.6 |
| >8589929.01 | 30.00 | 14.7 - 49.4 | 100.00 | 88.4 - 100.0 |  |  | 0.70 | 0.6 - 0.9 | 100.0 | 92.8 |
| >10054368.9199999 | 26.67 | 12.3 - 45.9 | 100.00 | 88.4 - 100.0 |  |  | 0.73 | 0.6 - 0.9 | 100.0 | 92.5 |
| >12574708.33 | 23.33 | 9.9 - 42.3 | 100.00 | 88.4 - 100.0 |  |  | 0.77 | 0.6 - 0.9 | 100.0 | 92.2 |
| >13270249.6199999 | 20.00 | 7.7 - 38.6 | 100.00 | 88.4 - 100.0 |  |  | 0.80 | 0.7 - 1.0 | 100.0 | 91.8 |
| >13594071.59 | 16.67 | 5.6 - 34.7 | 100.00 | 88.4 - 100.0 |  |  | 0.83 | 0.7 - 1.0 | 100.0 | 91.5 |
| >21333055.78 | 13.33 | 3.8 - 30.7 | 100.00 | 88.4 - 100.0 |  |  | 0.87 | 0.8 - 1.0 | 100.0 | 91.2 |
| >23286313.6799999 | 10.00 | 2.1 - 26.5 | 100.00 | 88.4 - 100.0 |  |  | 0.90 | 0.8 - 1.0 | 100.0 | 90.9 |
| >23350153.7599999 | 6.67 | 0.8 - 22.1 | 100.00 | 88.4 - 100.0 |  |  | 0.93 | 0.8 - 1.0 | 100.0 | 90.6 |
| >31210201.55 | 3.33 | 0.08 - 17.2 | 100.00 | 88.4 - 100.0 |  |  | 0.97 | 0.9 - 1.0 | 100.0 | 90.3 |
| >41731291.3999999 | 0.00 | 0.0 - 11.6 | 100.00 | 88.4 - 100.0 |  |  | 1.00 | 1.0 - 1.0 |  | 90.0 |

### ROC curve

| Variable | **78,600-80,500 m/z** |
| --- | --- |
| Classification variable | COVID-19_Status |

| Sample size | 60 |
| --- | --- |
| Positive group ^a^ | 30 (50.00%) |
| Negative group ^b^ | 30 (50.00%) |

^a^ COVID-19_Status = 1
^b^ COVID-19_Status = 0

| Disease prevalence (%) | 10 |
| --- | --- |

#### Area under the ROC curve (AUC)

| Area under the ROC curve (AUC) | 0.954 |
| --- | --- |
| Standard Error ^a^ | 0.0293 |
| 95% Confidence interval ^b^ | 0.867 to 0.991 |
| z statistic | 15.513 |
| Significance level P (Area=0.5) | <0.0001 |

^a^ DeLong et al., 1988

^b^ Binomial exact

#### Youden index

| Youden index J | 0.8333 |
| --- | --- |
| Associated criterion | >937903.91 |
| Sensitivity | 93.33 |
| Specificity | 90.00 |

#### Criterion values and coordinates of the ROC curve [[Show]](javascript:showdiv('d32','d33','table1');)

#### Criterion values and coordinates of the ROC curve

| Criterion | Sensitivity | 95% CI | Specificity | 95% CI | +LR | 95% CI | -LR | 95% CI | +PV | -PV |
| --- | --- | --- | --- | --- | --- | --- | --- | --- | --- | --- |
| ≥17750.38 | 100.00 | 88.4 - 100.0 | 0.00 | 0.0 - 11.6 | 1.00 | 1.0 - 1.0 |  |  | 10.0 |  |
| >17750.38 | 100.00 | 88.4 - 100.0 | 3.33 | 0.08 - 17.2 | 1.03 | 1.0 - 1.1 | 0.00 |  | 10.3 | 100.0 |
| >28537.38 | 100.00 | 88.4 - 100.0 | 6.67 | 0.8 - 22.1 | 1.07 | 1.0 - 1.2 | 0.00 |  | 10.6 | 100.0 |
| >28967.5 | 100.00 | 88.4 - 100.0 | 10.00 | 2.1 - 26.5 | 1.11 | 1.0 - 1.3 | 0.00 |  | 11.0 | 100.0 |
| >38065.78 | 100.00 | 88.4 - 100.0 | 13.33 | 3.8 - 30.7 | 1.15 | 1.0 - 1.3 | 0.00 |  | 11.4 | 100.0 |
| >39621.92 | 100.00 | 88.4 - 100.0 | 16.67 | 5.6 - 34.7 | 1.20 | 1.0 - 1.4 | 0.00 |  | 11.8 | 100.0 |
| >40305 | 100.00 | 88.4 - 100.0 | 20.00 | 7.7 - 38.6 | 1.25 | 1.0 - 1.5 | 0.00 |  | 12.2 | 100.0 |
| >53640.64 | 100.00 | 88.4 - 100.0 | 23.33 | 9.9 - 42.3 | 1.30 | 1.1 - 1.6 | 0.00 |  | 12.7 | 100.0 |
| >57099.11 | 100.00 | 88.4 - 100.0 | 26.67 | 12.3 - 45.9 | 1.36 | 1.1 - 1.7 | 0.00 |  | 13.2 | 100.0 |
| >83157.35 | 100.00 | 88.4 - 100.0 | 30.00 | 14.7 - 49.4 | 1.43 | 1.1 - 1.8 | 0.00 |  | 13.7 | 100.0 |
| >88345.87 | 100.00 | 88.4 - 100.0 | 33.33 | 17.3 - 52.8 | 1.50 | 1.2 - 1.9 | 0.00 |  | 14.3 | 100.0 |
| >90940.09 | 100.00 | 88.4 - 100.0 | 36.67 | 19.9 - 56.1 | 1.58 | 1.2 - 2.1 | 0.00 |  | 14.9 | 100.0 |
| >92744.88 | 100.00 | 88.4 - 100.0 | 40.00 | 22.7 - 59.4 | 1.67 | 1.2 - 2.2 | 0.00 |  | 15.6 | 100.0 |
| >170541.05 | 100.00 | 88.4 - 100.0 | 43.33 | 25.5 - 62.6 | 1.76 | 1.3 - 2.4 | 0.00 |  | 16.4 | 100.0 |
| >190049.15 | 100.00 | 88.4 - 100.0 | 46.67 | 28.3 - 65.7 | 1.87 | 1.3 - 2.6 | 0.00 |  | 17.2 | 100.0 |
| >202346.8 | 100.00 | 88.4 - 100.0 | 50.00 | 31.3 - 68.7 | 2.00 | 1.4 - 2.9 | 0.00 |  | 18.2 | 100.0 |
| >206213.99 | 100.00 | 88.4 - 100.0 | 53.33 | 34.3 - 71.7 | 2.14 | 1.5 - 3.1 | 0.00 |  | 19.2 | 100.0 |
| >228664.88 | 100.00 | 88.4 - 100.0 | 56.67 | 37.4 - 74.5 | 2.31 | 1.5 - 3.5 | 0.00 |  | 20.4 | 100.0 |
| >243896.21 | 100.00 | 88.4 - 100.0 | 60.00 | 40.6 - 77.3 | 2.50 | 1.6 - 3.9 | 0.00 |  | 21.7 | 100.0 |
| >280138.01 | 100.00 | 88.4 - 100.0 | 63.33 | 43.9 - 80.1 | 2.73 | 1.7 - 4.4 | 0.00 |  | 23.3 | 100.0 |
| >289542.32 | 100.00 | 88.4 - 100.0 | 66.67 | 47.2 - 82.7 | 3.00 | 1.8 - 5.0 | 0.00 |  | 25.0 | 100.0 |
| >332940.71 | 96.67 | 82.8 - 99.9 | 66.67 | 47.2 - 82.7 | 2.90 | 1.7 - 4.8 | 0.050 | 0.007 - 0.3 | 24.4 | 99.4 |
| >362696.06 | 96.67 | 82.8 - 99.9 | 70.00 | 50.6 - 85.3 | 3.22 | 1.9 - 5.6 | 0.048 | 0.007 - 0.3 | 26.4 | 99.5 |
| >381826.43 | 96.67 | 82.8 - 99.9 | 73.33 | 54.1 - 87.7 | 3.62 | 2.0 - 6.6 | 0.045 | 0.007 - 0.3 | 28.7 | 99.5 |
| >435651.24 | 96.67 | 82.8 - 99.9 | 76.67 | 57.7 - 90.1 | 4.14 | 2.2 - 8.0 | 0.043 | 0.006 - 0.3 | 31.5 | 99.5 |
| >573665.28 | 96.67 | 82.8 - 99.9 | 80.00 | 61.4 - 92.3 | 4.83 | 2.4 - 9.9 | 0.042 | 0.006 - 0.3 | 34.9 | 99.5 |
| >573910.05 | 96.67 | 82.8 - 99.9 | 83.33 | 65.3 - 94.4 | 5.80 | 2.6 - 12.9 | 0.040 | 0.006 - 0.3 | 39.2 | 99.6 |
| >666302.54 | 93.33 | 77.9 - 99.2 | 83.33 | 65.3 - 94.4 | 5.60 | 2.5 - 12.5 | 0.080 | 0.02 - 0.3 | 38.4 | 99.1 |
| >831384.93 | 93.33 | 77.9 - 99.2 | 86.67 | 69.3 - 96.2 | 7.00 | 2.8 - 17.5 | 0.077 | 0.02 - 0.3 | 43.8 | 99.2 |
| >937903.91 | 93.33 | 77.9 - 99.2 | 90.00 | 73.5 - 97.9 | 9.33 | 3.2 - 27.4 | 0.074 | 0.02 - 0.3 | 50.9 | 99.2 |
| >1269503.14 | 90.00 | 73.5 - 97.9 | 90.00 | 73.5 - 97.9 | 9.00 | 3.1 - 26.5 | 0.11 | 0.04 - 0.3 | 50.0 | 98.8 |
| >1408120 | 90.00 | 73.5 - 97.9 | 93.33 | 77.9 - 99.2 | 13.50 | 3.5 - 51.8 | 0.11 | 0.04 - 0.3 | 60.0 | 98.8 |
| >1480673.91 | 86.67 | 69.3 - 96.2 | 93.33 | 77.9 - 99.2 | 13.00 | 3.4 - 50.0 | 0.14 | 0.06 - 0.4 | 59.1 | 98.4 |
| >1504065.24 | 83.33 | 65.3 - 94.4 | 93.33 | 77.9 - 99.2 | 12.50 | 3.2 - 48.1 | 0.18 | 0.08 - 0.4 | 58.1 | 98.1 |
| >1557519.82 | 83.33 | 65.3 - 94.4 | 96.67 | 82.8 - 99.9 | 25.00 | 3.6 - 172.9 | 0.17 | 0.08 - 0.4 | 73.5 | 98.1 |
| >1668702.55 | 80.00 | 61.4 - 92.3 | 96.67 | 82.8 - 99.9 | 24.00 | 3.5 - 166.2 | 0.21 | 0.1 - 0.4 | 72.7 | 97.8 |
| >1711378.9 | 76.67 | 57.7 - 90.1 | 96.67 | 82.8 - 99.9 | 23.00 | 3.3 - 159.6 | 0.24 | 0.1 - 0.5 | 71.9 | 97.4 |
| >1728302.38 | 73.33 | 54.1 - 87.7 | 96.67 | 82.8 - 99.9 | 22.00 | 3.2 - 153.0 | 0.28 | 0.2 - 0.5 | 71.0 | 97.0 |
| >1748690.04 | 70.00 | 50.6 - 85.3 | 96.67 | 82.8 - 99.9 | 21.00 | 3.0 - 146.3 | 0.31 | 0.2 - 0.5 | 70.0 | 96.7 |
| >1835468.93 | 66.67 | 47.2 - 82.7 | 96.67 | 82.8 - 99.9 | 20.00 | 2.9 - 139.7 | 0.34 | 0.2 - 0.6 | 69.0 | 96.3 |
| >1925542.32 | 63.33 | 43.9 - 80.1 | 96.67 | 82.8 - 99.9 | 19.00 | 2.7 - 133.0 | 0.38 | 0.2 - 0.6 | 67.9 | 96.0 |
| >1954662.57 | 60.00 | 40.6 - 77.3 | 96.67 | 82.8 - 99.9 | 18.00 | 2.6 - 126.4 | 0.41 | 0.3 - 0.6 | 66.7 | 95.6 |
| >2057250.62 | 56.67 | 37.4 - 74.5 | 96.67 | 82.8 - 99.9 | 17.00 | 2.4 - 119.8 | 0.45 | 0.3 - 0.7 | 65.4 | 95.3 |
| >2109966.2 | 53.33 | 34.3 - 71.7 | 96.67 | 82.8 - 99.9 | 16.00 | 2.3 - 113.1 | 0.48 | 0.3 - 0.7 | 64.0 | 94.9 |
| >2183127.46 | 50.00 | 31.3 - 68.7 | 96.67 | 82.8 - 99.9 | 15.00 | 2.1 - 106.5 | 0.52 | 0.4 - 0.7 | 62.5 | 94.6 |
| >3097754.66 | 46.67 | 28.3 - 65.7 | 96.67 | 82.8 - 99.9 | 14.00 | 2.0 - 99.9 | 0.55 | 0.4 - 0.8 | 60.9 | 94.2 |
| >3159487.61 | 43.33 | 25.5 - 62.6 | 96.67 | 82.8 - 99.9 | 13.00 | 1.8 - 93.2 | 0.59 | 0.4 - 0.8 | 59.1 | 93.9 |
| >3224982.16 | 40.00 | 22.7 - 59.4 | 96.67 | 82.8 - 99.9 | 12.00 | 1.7 - 86.6 | 0.62 | 0.5 - 0.8 | 57.1 | 93.5 |
| >3331148.94 | 36.67 | 19.9 - 56.1 | 96.67 | 82.8 - 99.9 | 11.00 | 1.5 - 80.0 | 0.66 | 0.5 - 0.9 | 55.0 | 93.2 |
| >3331823.28 | 33.33 | 17.3 - 52.8 | 96.67 | 82.8 - 99.9 | 10.00 | 1.4 - 73.3 | 0.69 | 0.5 - 0.9 | 52.6 | 92.9 |
| >3639969.6 | 30.00 | 14.7 - 49.4 | 96.67 | 82.8 - 99.9 | 9.00 | 1.2 - 66.7 | 0.72 | 0.6 - 0.9 | 50.0 | 92.6 |
| >4611529.82 | 26.67 | 12.3 - 45.9 | 96.67 | 82.8 - 99.9 | 8.00 | 1.1 - 60.1 | 0.76 | 0.6 - 1.0 | 47.1 | 92.2 |
| >5326420.86 | 23.33 | 9.9 - 42.3 | 96.67 | 82.8 - 99.9 | 7.00 | 0.9 - 53.5 | 0.79 | 0.6 - 1.0 | 43.8 | 91.9 |
| >6009479.37 | 20.00 | 7.7 - 38.6 | 96.67 | 82.8 - 99.9 | 6.00 | 0.8 - 46.9 | 0.83 | 0.7 - 1.0 | 40.0 | 91.6 |
| >6437387.04999999 | 20.00 | 7.7 - 38.6 | 100.00 | 88.4 - 100.0 |  |  | 0.80 | 0.7 - 1.0 | 100.0 | 91.8 |
| >7520075.97 | 16.67 | 5.6 - 34.7 | 100.00 | 88.4 - 100.0 |  |  | 0.83 | 0.7 - 1.0 | 100.0 | 91.5 |
| >8380270.1 | 13.33 | 3.8 - 30.7 | 100.00 | 88.4 - 100.0 |  |  | 0.87 | 0.8 - 1.0 | 100.0 | 91.2 |
| >8742368.28999999 | 10.00 | 2.1 - 26.5 | 100.00 | 88.4 - 100.0 |  |  | 0.90 | 0.8 - 1.0 | 100.0 | 90.9 |
| >10780855.5999999 | 6.67 | 0.8 - 22.1 | 100.00 | 88.4 - 100.0 |  |  | 0.93 | 0.8 - 1.0 | 100.0 | 90.6 |
| >15569879.47 | 3.33 | 0.08 - 17.2 | 100.00 | 88.4 - 100.0 |  |  | 0.97 | 0.9 - 1.0 | 100.0 | 90.3 |
| >30283238.8699999 | 0.00 | 0.0 - 11.6 | 100.00 | 88.4 - 100.0 |  |  | 1.00 | 1.0 - 1.0 |  | 90.0 |

### ROC curve

| Variable | **111,500-115,500 m/z** |
| --- | --- |
| Classification variable | COVID-19_Status |

| Sample size | 60 |
| --- | --- |
| Positive group ^a^ | 30 (50.00%) |
| Negative group ^b^ | 30 (50.00%) |

^a^ COVID-19_Status = 1
^b^ COVID-19_Status = 0

| Disease prevalence (%) | 10 |
| --- | --- |

#### Area under the ROC curve (AUC)

| Area under the ROC curve (AUC) | 0.933 |
| --- | --- |
| Standard Error ^a^ | 0.0355 |
| 95% Confidence interval ^b^ | 0.838 to 0.982 |
| z statistic | 12.211 |
| Significance level P (Area=0.5) | <0.0001 |

^a^ DeLong et al., 1988

^b^ Binomial exact

#### Youden index

| Youden index J | 0.8333 |
| --- | --- |
| Associated criterion | >1104743.08 |
| Sensitivity | 93.33 |
| Specificity | 90.00 |

#### Criterion values and coordinates of the ROC curve [[Show]](javascript:showdiv('d34','d35','table1');)

#### Criterion values and coordinates of the ROC curve

| Criterion | Sensitivity | 95% CI | Specificity | 95% CI | +LR | 95% CI | -LR | 95% CI | +PV | -PV |
| --- | --- | --- | --- | --- | --- | --- | --- | --- | --- | --- |
| ≥18023.67 | 100.00 | 88.4 - 100.0 | 0.00 | 0.0 - 11.6 | 1.00 | 1.0 - 1.0 |  |  | 10.0 |  |
| >18023.67 | 100.00 | 88.4 - 100.0 | 3.33 | 0.08 - 17.2 | 1.03 | 1.0 - 1.1 | 0.00 |  | 10.3 | 100.0 |
| >20702.83 | 100.00 | 88.4 - 100.0 | 6.67 | 0.8 - 22.1 | 1.07 | 1.0 - 1.2 | 0.00 |  | 10.6 | 100.0 |
| >23752.02 | 100.00 | 88.4 - 100.0 | 10.00 | 2.1 - 26.5 | 1.11 | 1.0 - 1.3 | 0.00 |  | 11.0 | 100.0 |
| >25461.24 | 100.00 | 88.4 - 100.0 | 13.33 | 3.8 - 30.7 | 1.15 | 1.0 - 1.3 | 0.00 |  | 11.4 | 100.0 |
| >36732.2 | 100.00 | 88.4 - 100.0 | 16.67 | 5.6 - 34.7 | 1.20 | 1.0 - 1.4 | 0.00 |  | 11.8 | 100.0 |
| >46684.3 | 100.00 | 88.4 - 100.0 | 20.00 | 7.7 - 38.6 | 1.25 | 1.0 - 1.5 | 0.00 |  | 12.2 | 100.0 |
| >54864.46 | 100.00 | 88.4 - 100.0 | 23.33 | 9.9 - 42.3 | 1.30 | 1.1 - 1.6 | 0.00 |  | 12.7 | 100.0 |
| >62801.6 | 100.00 | 88.4 - 100.0 | 26.67 | 12.3 - 45.9 | 1.36 | 1.1 - 1.7 | 0.00 |  | 13.2 | 100.0 |
| >70848.77 | 100.00 | 88.4 - 100.0 | 30.00 | 14.7 - 49.4 | 1.43 | 1.1 - 1.8 | 0.00 |  | 13.7 | 100.0 |
| >85216.56 | 100.00 | 88.4 - 100.0 | 33.33 | 17.3 - 52.8 | 1.50 | 1.2 - 1.9 | 0.00 |  | 14.3 | 100.0 |
| >85942.51 | 100.00 | 88.4 - 100.0 | 36.67 | 19.9 - 56.1 | 1.58 | 1.2 - 2.1 | 0.00 |  | 14.9 | 100.0 |
| >110193.24 | 100.00 | 88.4 - 100.0 | 40.00 | 22.7 - 59.4 | 1.67 | 1.2 - 2.2 | 0.00 |  | 15.6 | 100.0 |
| >117172.46 | 96.67 | 82.8 - 99.9 | 40.00 | 22.7 - 59.4 | 1.61 | 1.2 - 2.2 | 0.083 | 0.01 - 0.6 | 15.2 | 99.1 |
| >154924.29 | 96.67 | 82.8 - 99.9 | 43.33 | 25.5 - 62.6 | 1.71 | 1.2 - 2.3 | 0.077 | 0.01 - 0.6 | 15.9 | 99.2 |
| >167513.37 | 96.67 | 82.8 - 99.9 | 46.67 | 28.3 - 65.7 | 1.81 | 1.3 - 2.5 | 0.071 | 0.01 - 0.5 | 16.8 | 99.2 |
| >199242.98 | 96.67 | 82.8 - 99.9 | 50.00 | 31.3 - 68.7 | 1.93 | 1.3 - 2.8 | 0.067 | 0.009 - 0.5 | 17.7 | 99.3 |
| >201059.48 | 96.67 | 82.8 - 99.9 | 53.33 | 34.3 - 71.7 | 2.07 | 1.4 - 3.1 | 0.063 | 0.009 - 0.4 | 18.7 | 99.3 |
| >214531.44 | 96.67 | 82.8 - 99.9 | 56.67 | 37.4 - 74.5 | 2.23 | 1.5 - 3.4 | 0.059 | 0.008 - 0.4 | 19.9 | 99.4 |
| >257348.53 | 96.67 | 82.8 - 99.9 | 60.00 | 40.6 - 77.3 | 2.42 | 1.6 - 3.8 | 0.056 | 0.008 - 0.4 | 21.2 | 99.4 |
| >269446.62 | 96.67 | 82.8 - 99.9 | 63.33 | 43.9 - 80.1 | 2.64 | 1.6 - 4.2 | 0.053 | 0.008 - 0.4 | 22.7 | 99.4 |
| >288291.2 | 96.67 | 82.8 - 99.9 | 66.67 | 47.2 - 82.7 | 2.90 | 1.7 - 4.8 | 0.050 | 0.007 - 0.3 | 24.4 | 99.4 |
| >351605.87 | 96.67 | 82.8 - 99.9 | 70.00 | 50.6 - 85.3 | 3.22 | 1.9 - 5.6 | 0.048 | 0.007 - 0.3 | 26.4 | 99.5 |
| >385289.14 | 96.67 | 82.8 - 99.9 | 73.33 | 54.1 - 87.7 | 3.62 | 2.0 - 6.6 | 0.045 | 0.007 - 0.3 | 28.7 | 99.5 |
| >398905.76 | 96.67 | 82.8 - 99.9 | 76.67 | 57.7 - 90.1 | 4.14 | 2.2 - 8.0 | 0.043 | 0.006 - 0.3 | 31.5 | 99.5 |
| >737500.85 | 96.67 | 82.8 - 99.9 | 80.00 | 61.4 - 92.3 | 4.83 | 2.4 - 9.9 | 0.042 | 0.006 - 0.3 | 34.9 | 99.5 |
| >767758.8 | 96.67 | 82.8 - 99.9 | 83.33 | 65.3 - 94.4 | 5.80 | 2.6 - 12.9 | 0.040 | 0.006 - 0.3 | 39.2 | 99.6 |
| >935163.35 | 93.33 | 77.9 - 99.2 | 83.33 | 65.3 - 94.4 | 5.60 | 2.5 - 12.5 | 0.080 | 0.02 - 0.3 | 38.4 | 99.1 |
| >1059039.66 | 93.33 | 77.9 - 99.2 | 86.67 | 69.3 - 96.2 | 7.00 | 2.8 - 17.5 | 0.077 | 0.02 - 0.3 | 43.8 | 99.2 |
| >1104743.08 | 93.33 | 77.9 - 99.2 | 90.00 | 73.5 - 97.9 | 9.33 | 3.2 - 27.4 | 0.074 | 0.02 - 0.3 | 50.9 | 99.2 |
| >1284275.03 | 90.00 | 73.5 - 97.9 | 90.00 | 73.5 - 97.9 | 9.00 | 3.1 - 26.5 | 0.11 | 0.04 - 0.3 | 50.0 | 98.8 |
| >1396596.27 | 86.67 | 69.3 - 96.2 | 90.00 | 73.5 - 97.9 | 8.67 | 2.9 - 25.6 | 0.15 | 0.06 - 0.4 | 49.1 | 98.4 |
| >1764364.59 | 86.67 | 69.3 - 96.2 | 93.33 | 77.9 - 99.2 | 13.00 | 3.4 - 50.0 | 0.14 | 0.06 - 0.4 | 59.1 | 98.4 |
| >1851531.23 | 83.33 | 65.3 - 94.4 | 93.33 | 77.9 - 99.2 | 12.50 | 3.2 - 48.1 | 0.18 | 0.08 - 0.4 | 58.1 | 98.1 |
| >2056313.79 | 80.00 | 61.4 - 92.3 | 93.33 | 77.9 - 99.2 | 12.00 | 3.1 - 46.3 | 0.21 | 0.1 - 0.4 | 57.1 | 97.7 |
| >2123447.29 | 76.67 | 57.7 - 90.1 | 93.33 | 77.9 - 99.2 | 11.50 | 3.0 - 44.5 | 0.25 | 0.1 - 0.5 | 56.1 | 97.3 |
| >2478022.46 | 73.33 | 54.1 - 87.7 | 93.33 | 77.9 - 99.2 | 11.00 | 2.8 - 42.7 | 0.29 | 0.2 - 0.5 | 55.0 | 96.9 |
| >2858718.17 | 70.00 | 50.6 - 85.3 | 93.33 | 77.9 - 99.2 | 10.50 | 2.7 - 40.9 | 0.32 | 0.2 - 0.6 | 53.8 | 96.6 |
| >2872345.82 | 66.67 | 47.2 - 82.7 | 93.33 | 77.9 - 99.2 | 10.00 | 2.6 - 39.1 | 0.36 | 0.2 - 0.6 | 52.6 | 96.2 |
| >3823806.79 | 63.33 | 43.9 - 80.1 | 93.33 | 77.9 - 99.2 | 9.50 | 2.4 - 37.2 | 0.39 | 0.2 - 0.6 | 51.4 | 95.8 |
| >4278858.73 | 60.00 | 40.6 - 77.3 | 93.33 | 77.9 - 99.2 | 9.00 | 2.3 - 35.4 | 0.43 | 0.3 - 0.7 | 50.0 | 95.5 |
| >4389825.29 | 56.67 | 37.4 - 74.5 | 93.33 | 77.9 - 99.2 | 8.50 | 2.1 - 33.6 | 0.46 | 0.3 - 0.7 | 48.6 | 95.1 |
| >5153960.79 | 53.33 | 34.3 - 71.7 | 93.33 | 77.9 - 99.2 | 8.00 | 2.0 - 31.8 | 0.50 | 0.3 - 0.7 | 47.1 | 94.7 |
| >5991863.27999999 | 50.00 | 31.3 - 68.7 | 93.33 | 77.9 - 99.2 | 7.50 | 1.9 - 30.0 | 0.54 | 0.4 - 0.8 | 45.5 | 94.4 |
| >6186171.84 | 46.67 | 28.3 - 65.7 | 93.33 | 77.9 - 99.2 | 7.00 | 1.7 - 28.2 | 0.57 | 0.4 - 0.8 | 43.8 | 94.0 |
| >6813772.24999999 | 43.33 | 25.5 - 62.6 | 93.33 | 77.9 - 99.2 | 6.50 | 1.6 - 26.4 | 0.61 | 0.4 - 0.8 | 41.9 | 93.7 |
| >8336401.97 | 40.00 | 22.7 - 59.4 | 93.33 | 77.9 - 99.2 | 6.00 | 1.5 - 24.5 | 0.64 | 0.5 - 0.9 | 40.0 | 93.3 |
| >8982387.57 | 36.67 | 19.9 - 56.1 | 93.33 | 77.9 - 99.2 | 5.50 | 1.3 - 22.7 | 0.68 | 0.5 - 0.9 | 37.9 | 93.0 |
| >9121424.11999999 | 36.67 | 19.9 - 56.1 | 96.67 | 82.8 - 99.9 | 11.00 | 1.5 - 80.0 | 0.66 | 0.5 - 0.9 | 55.0 | 93.2 |
| >9361286.42 | 33.33 | 17.3 - 52.8 | 96.67 | 82.8 - 99.9 | 10.00 | 1.4 - 73.3 | 0.69 | 0.5 - 0.9 | 52.6 | 92.9 |
| >9419206.24999999 | 33.33 | 17.3 - 52.8 | 100.00 | 88.4 - 100.0 |  |  | 0.67 | 0.5 - 0.9 | 100.0 | 93.1 |
| >11508762.9899999 | 30.00 | 14.7 - 49.4 | 100.00 | 88.4 - 100.0 |  |  | 0.70 | 0.6 - 0.9 | 100.0 | 92.8 |
| >13790682.8099999 | 26.67 | 12.3 - 45.9 | 100.00 | 88.4 - 100.0 |  |  | 0.73 | 0.6 - 0.9 | 100.0 | 92.5 |
| >16898158.01 | 23.33 | 9.9 - 42.3 | 100.00 | 88.4 - 100.0 |  |  | 0.77 | 0.6 - 0.9 | 100.0 | 92.2 |
| >18349577.5999999 | 20.00 | 7.7 - 38.6 | 100.00 | 88.4 - 100.0 |  |  | 0.80 | 0.7 - 1.0 | 100.0 | 91.8 |
| >20891101.0299999 | 16.67 | 5.6 - 34.7 | 100.00 | 88.4 - 100.0 |  |  | 0.83 | 0.7 - 1.0 | 100.0 | 91.5 |
| >25307092.4699999 | 13.33 | 3.8 - 30.7 | 100.00 | 88.4 - 100.0 |  |  | 0.87 | 0.8 - 1.0 | 100.0 | 91.2 |
| >33089507.49 | 10.00 | 2.1 - 26.5 | 100.00 | 88.4 - 100.0 |  |  | 0.90 | 0.8 - 1.0 | 100.0 | 90.9 |
| >74857005.1899999 | 6.67 | 0.8 - 22.1 | 100.00 | 88.4 - 100.0 |  |  | 0.93 | 0.8 - 1.0 | 100.0 | 90.6 |
| >80162113.5499999 | 3.33 | 0.08 - 17.2 | 100.00 | 88.4 - 100.0 |  |  | 0.97 | 0.9 - 1.0 | 100.0 | 90.3 |
| >130894046.4 | 0.00 | 0.0 - 11.6 | 100.00 | 88.4 - 100.0 |  |  | 1.00 | 1.0 - 1.0 |  | 90.0 |
